# Quantifying proximity, confinement, and interventions in disease outbreaks: a decision support framework for air-transported pathogens

**DOI:** 10.1101/2020.09.09.20187625

**Authors:** Tami C. Bond, Angela Bosco-Lauth, Delphine K. Farmer, Paul W. Francisco, Jeffrey R. Pierce, Kristen M. Fedak, Jay M. Ham, Shantanu H. Jathar, Sue VandeWoude

## Abstract

The inability to communicate how infectious diseases are transmitted in human environments has triggered avoidance of interactions during the COVID-19 pandemic. We define a metric, Effective ReBreathed Volume (ERBV), that encapsulates how infectious pathogens transport in air. This measure distinguishes environmental transport from other factors in the chain of infection, thus allowing quantitative comparisons of the riskiness of different situations for any pathogens transported in air, including SARS-CoV-2. Particle size is a key factor in transport, removal onto surfaces, and elimination by mitigation measures, so ERBV is presented for a range of exhaled particle diameters: 1 μm, 10 μm, and 100 μm. Pathogen transport is enhanced by two separate but interacting effects: proximity and confinement. Confinement in enclosed spaces overwhelms proximity after 10–15 minutes for all but the largest particles. Therefore, we review plausible strategies to reduce the confinement effect. Changes in standard ventilation and filtration can reduce person-to-person transport of 1-μm particles (ERBV_1_) by 13-85% in residential and commercial situations. Deposition to surfaces competes with intentional removal for 10-μm and 100-μm particles, so the same interventions reduce ERBV_10_ by only 3-50%, and ERBV_100_ is unaffected. Determining transmission modes is critical to identify intervention effectiveness, and would be accelerated with prior knowledge of ERBV. When judiciously selected, the interventions examined can provide substantial reduction in risk, and the conditions for selection are identified. The framework of size-dependent ERBV supports analysis and mitigation decisions in an emerging situation, even before other infectious parameters are well known.

## Introduction

The spread of the SARS-CoV-2 virus has created a public health crisis and widespread economic disruption (1). Key factors in the extent of this crisis are (i) the severity of the disease, COVID-19, so avoidance is preferred over illness; (ii) transmission by asymptomatic or presymptomatic individuals (2, 3); and (iii) the novelty of the disease, so that decisions must occur before scientific investigations are definitive. Although this situation is unprecedented in the past century, pandemics have occurred throughout human history. An event like COVID-19 was predicted before its onset (4) and is likely to occur again with different infection dynamics (5, 6).

Figure 1 illustrates the chain of infection (7), modified to emphasize the role of person-to-person interactions. After a pathogen has entered the human population, escape from the human reservoir depends on the prevalence and characteristics of disease carriers or emitters. On the receiving end, the likelihood of infection is determined by the host’s susceptibility and the dose received. The mode is the method of travel between the pathogen’s release and the host. The pathogen’s survival characteristics limit viable modes, but the environment modulates the transferred dose. This environment includes the social system that compels intersection between individuals, and the physical environment through which the pathogen travels. Uncertainty about physical transmission has led to suspicion about the interactions that underpin the economy. The ability to quantify exposure risks in social interactions more quickly and rigorously would aid decision-making in current and future outbreak situations.

Describing the chain of infection requires expertise in epidemiology, infectious disease, sociology and data science, engineered and natural environments, virology, immunology, and public health. Each field has burgeoned since Riley’s pioneering work (8) combined carrier and environmental characteristics into a single equation, yet few metrics distill the essential elements of the chain for use in collaboration. This paper describes a metric to quantify pathogen transport and uses it to compare transmission environments and mitigation measures.

**Figure 1.**
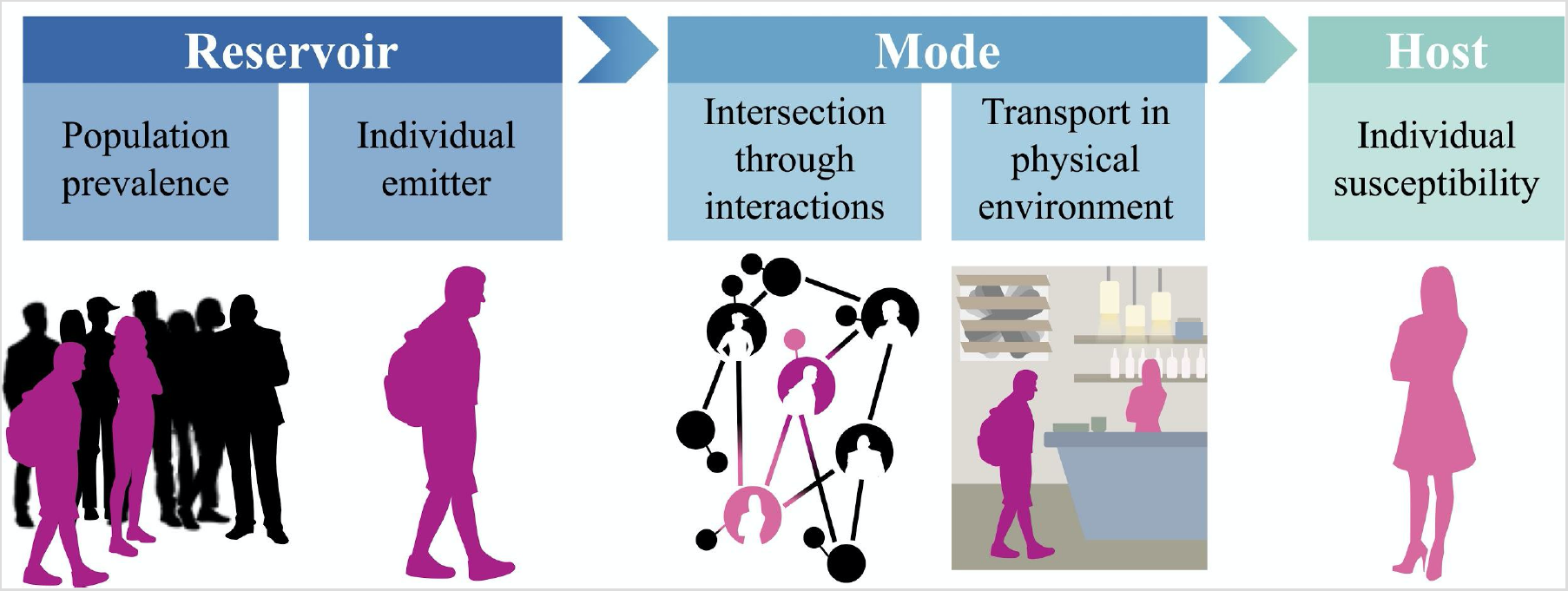
Chain of infection for a disease whose spread depends on human interactions. Figure credit: Mj Riches

### Quantifying person-to-person transport

A metric to characterize and communicate person-to-person transport would be understandable by individuals outside the field; able to encapsulate complex situations and incorporate evolving knowledge; generalizable to archetypal building situations; and germane to decision-making by comparing alternative interactions. It should not be confounded by differences in human emitters or recipients, which are independent of transport. We choose *rebreathed volume* (RBV) as a basic metric for this purpose. RBV is the total volume of air exhaled by one person and subsequently inhaled by another. RBV is proportional to the *total* dose that an individual receives; we also use the *rate* of rebreathing to compare different interactions of equal lengths. If a recipient were inhaling directly from the mouth of an emitter, the rebreathing rate would be 8 L min^−1^, and over 10 minutes, RBV would be 80 L. RBV can be calculated from simple models, computational fluid dynamic models, and tracer measurements in both indoor and outdoor situations (S.1-S.4). RBV is similar to other metrics and can be calculated from them (S.10), including the Wells-Riley equation for probability of infection (8), inhalation intake fraction (9), or rebreathed fraction (10). Because RBV quantifies person-to-person transmission, the number of emitters also needs to be included in risk of infection. An additional “crowding” effect should be calculated separately.

A particular challenge in any emerging situation is uncertainty in the mode of transmission. Public health guidance (11) uses the terms “droplet” and “short-range” for large expiratory particles that transport through air but are lost quickly by falling. A second mode, via small particles that tend to follow airstreams, is termed “airborne,” “aerosol,” or “long-range.” A third mode is called “indirect” when pathogens are transferred through intermediate, contaminated objects, including human skin (12). Dominant modes of transmission are hotly debated for COVID-19 (13, 14) and other respiratory infections (15).

Despite the differences in terminology, the dynamics of transport through air govern the first two modes, and play a role in the third. Our approach does not champion any particular mode, but instead acknowledges the importance of particle size in every step of the chain of infection. Particle size and viral content is influenced by where particles originate within the respiratory tract (16); size affects the depth of penetration into the recipient’s lungs and susceptibility (17). Size dominates particle fate; large particles do not remain suspended as long and are easier to remove because of the relative influences of gravity, drag force, and attachment to surfaces or deposition.

To communicate transport dynamics of differently sized particles while maintaining simplicity, we define *effective rebreathed volume* (ERBV) as the exhaled volume that contains the same number of particles as the air inhaled by the recipient. If a recipient received 80 L of RBV from an emitter, and 90% of particles with diameter X were lost by settling, then ERBV_X_ would be 8 L (80 L multiplied by 10% remaining). This physics-based treatment allows objective comparison of modes by accounting for the main difference in particle transport: size-dependent loss.

We choose decadally-spaced sizes that cover a biologically-relevant range: 1, 10 and 100 μm (ERBV_1_, ERBV_10_, and ERBV_100_, respectively). Diameters of expiratory particles range from 0.01 to 1000 μm (18–20), although the largest sizes are rarely measured. Particles the size of a bare virion (0.1–0.2 µm particles because they have similar indoor deposition loss rates (SI). 1000-μm particles are excluded because they would travel less than 1 meter due to their rapid fall speeds. Large expiratory droplets evaporate within a few seconds (21, 22), and a 100-μm droplet would become about 20 μm after losing 99% water content (23).

### Proximity and confinement effects

Person-to-person transport of pathogens is greater in close *proximity*, partly because contaminants spread out (disperse) as they travel away from an emitter, and partly because they also fall out (deposit) during that travel. Person-to-person transport is also greater in close *confinement*, where contaminants accumulate when they cannot escape the walls of an enclosure. Ventilation and other removal processes, including deposition, decrease the rate of accumulation in the confined space. Thus, the dose transferred from an emitter to a recipient depends upon dispersion, deposition, and other removal processes that lessen accumulation. Figure 2 compares the rate of rebreathing during simple maximum outdoor (red, shaded) and indoor (blue, dashed) interactions for 1-μm, 10-μm, and 100-μm particles. The contrast between the three particle sizes shows the importance of separate consideration.

**Figure 2.**
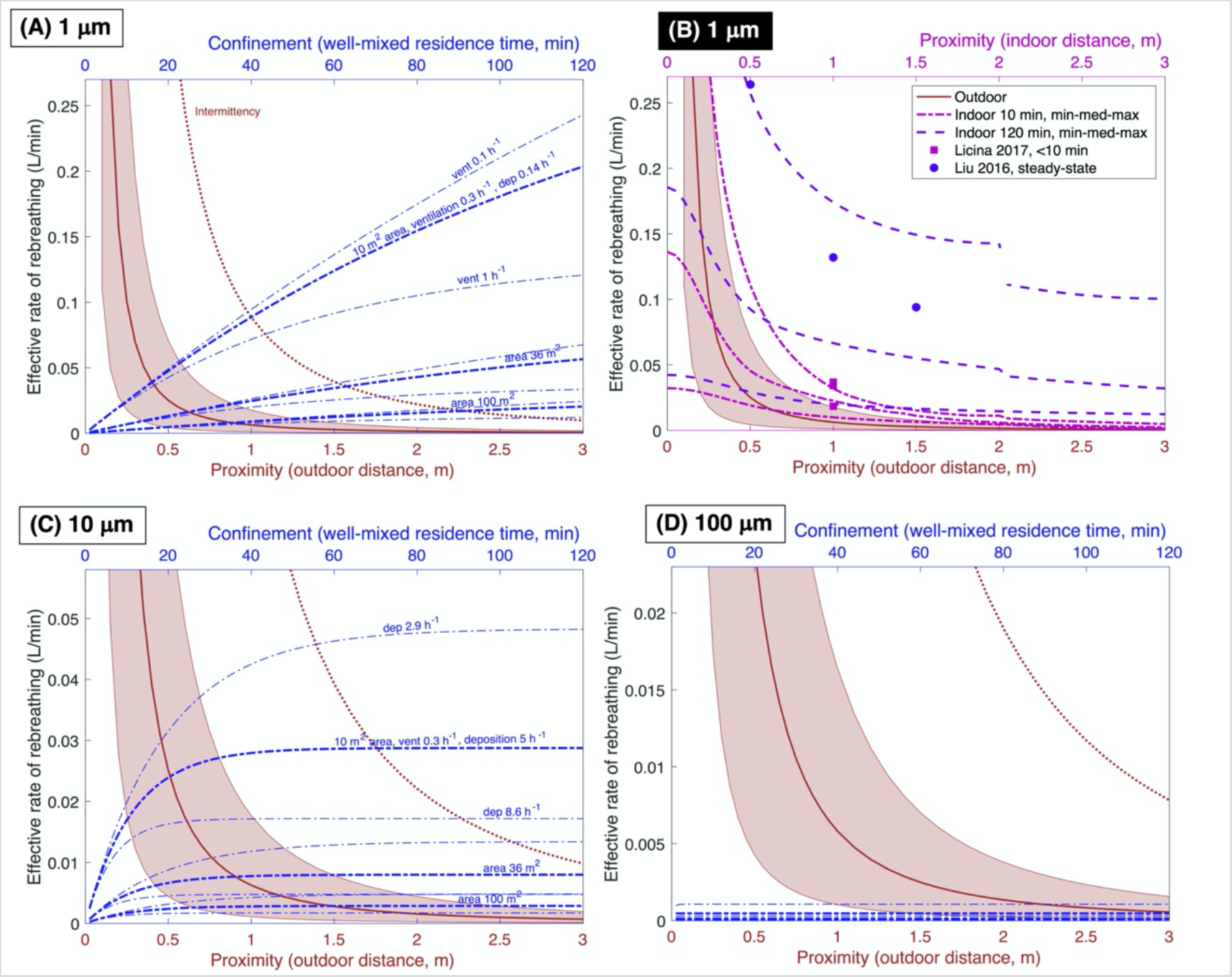
*Comparison of instantaneous effective rebreathing rate for outdoor (red, with shaded area) and indoor (blue, dashed) interactions for particles of diameter (A) 1 μm; (C) 10 μm; (D) 100 μm. Proximity (lower) and confinement (upper) axes are not equivalent, but appear on the same figure for comparison. Outdoors, ERBV depends on proximity. Shaded region shows uncertainty in atmospheric dispersion with wind at 2 m s^−1^; intermittency (dotted red line) is the estimated maximum value due to emission-plume meandering. Indoors in a well-mixed room, confinement affects ERBV and depends on the time of accumulation. Thick blue dashed lines show a range of room sizes. Ventilation rates affect rebreathing for 1-μm particles (light blue lines in [A]) and not for 100-μm particles (D); rates of deposition to surfaces affect rebreathing for 10-μm particles (C), as do ventilation rates (not shown). Panel (B) compares outdoor and indoor proximity for a range of simulated indoor conditions, for 1-μm particles at 10 min and 120 min after emitter entry. Highest ERBV occurs in small rooms, which disappear from the summary curve when the maximum possible distance from emitter is reached, creating the discontinuity shown in the dashed lines. Rebreathing rates are person-to-person and would increase for more emitters*.

Outside buildings, concentration decreases with distance from the emitter, because emitted particles are carried by wind and dispersed by air fluctuations. The proximity effect is especially attributable to dispersion, but also deposition for large particles. The gravitational settling that differentiates particles is negligible for 1-μm and 10-μm particles, but some 100-μm particles have fallen out after traveling 2–3 m. Public guidance in 2020 suggests maintaining 2-m separation between individuals, avoiding the highest concentrations. At this distance, the outdoor rebreathing rate is less than 0.01 L min^−1^, and would be even lower if the recipient were not directly downwind (Table 1).

**Table 1.**
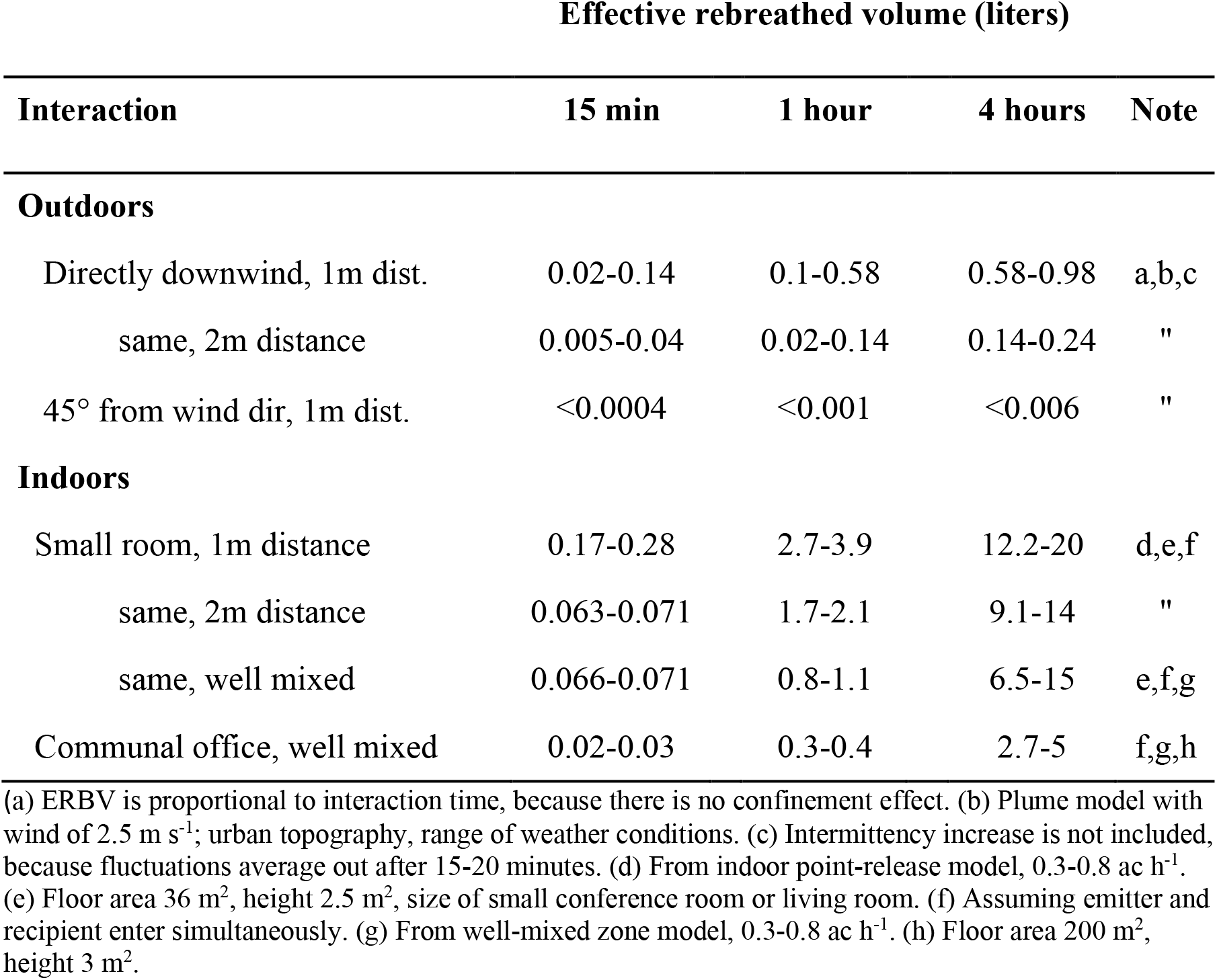
Person-to-person effective rebreathed volume for 1-µm particles (ERBV_1_) in common interactions.

Figure 2 also shows rebreathing in well-mixed, enclosed rooms (blue dashed curves), where the confinement effect occurs because exhaled air accumulates rather than dispersing. The rate of rebreathing depends on the room size, removal rates, and length of accumulation. Within 15 minutes indoors, the rebreathing rate for 1-µm and 10-µm particles exceeds that of a 2-m distance outdoors, with more rebreathing in small rooms. An individual who is unwilling to stand within 2 m of a potential emitter outdoors may unwittingly accept the same or greater risk by remaining in a moderate-sized room for 15 minutes.

The confinement effect is ameliorated by particle losses, which occur as particle-laden air travels out of the room through building cracks or through mechanical ventilation systems, or as they deposit on surfaces (24). Ventilation reduces the rebreathing rate noticeably for interaction times above about 30 minutes (Fig 2A). Particles of 100-µm deposit rapidly and do not accrue, so indoor rebreathing is low, consistent with particles classically termed “droplets” (Fig 2D). Particles of 10 μm diameter deposit more quickly than smaller particles, so indoor values of ERBV_10_ tend to be lower than ERBV_1_ (Fig 2B). Nevertheless, rebreathing of 10-μm particles is still noticeable in confined spaces, and these particles may be the residue of larger evaporated particles.

Both proximity and confinement effects occur in enclosed spaces. Contaminants disperse quickly indoors until they are well mixed (25). Figure 2B shows indoor rebreathing rates simulated with a point-release model (26, 27) for a range of simplified dispersion rates (SI). After 10 minutes of residence time, indoor and outdoor proximity effects are similar. An elevated proximity effect, as well as a confinement effect throughout the room, occur after two hours. Indoor dispersion rates depend on the intensity of turbulence in the room, which in turn is affected by environmental conditions, surface properties, and sources of thermal energy. Even the buoyancy around a human body can affect dispersion (28). Also shown are values interpreted from chamber measurements (28, 29). Both simulations and measurements can quantify rebreathing in specific situations, but precise estimates are not needed for effective guidance to address both proximity and confinement.

Total ERBV is obtained by summing over the recipient’s entire residence time. Table 1 summarizes ERBV_1_ over 15-minute, 1-hour, and 4-hour interactions, which represent a brief face-to-face commercial transaction, a business meeting, and a half-day working session, respectively. The definition of “close contact” from the Centers for Disease Control and Prevention is “within 6 feet for at least 15 minutes,” corresponding to a minimum ERBV of about 0.07 L for any particle size. Regardless of whether the participants are farther apart than 2-m distance, confinement in the two smaller rooms in Fig. 2 causes ERBV to exceed the “close-contact” value after about 10-15 minutes, for both 1-µm and 10-μm particles.

## Effect of mitigation measures

Rebreathing can be lessened when the participants remain at a distance to reduce proximity effects. Other solutions are needed to reduce the confinement effect, and those are explored here. Figure 3 summarizes ERBV for a 4-hour stay in residential (Fig 3A) and commercial (Fig. 3B) settings.

**Figure 3.**
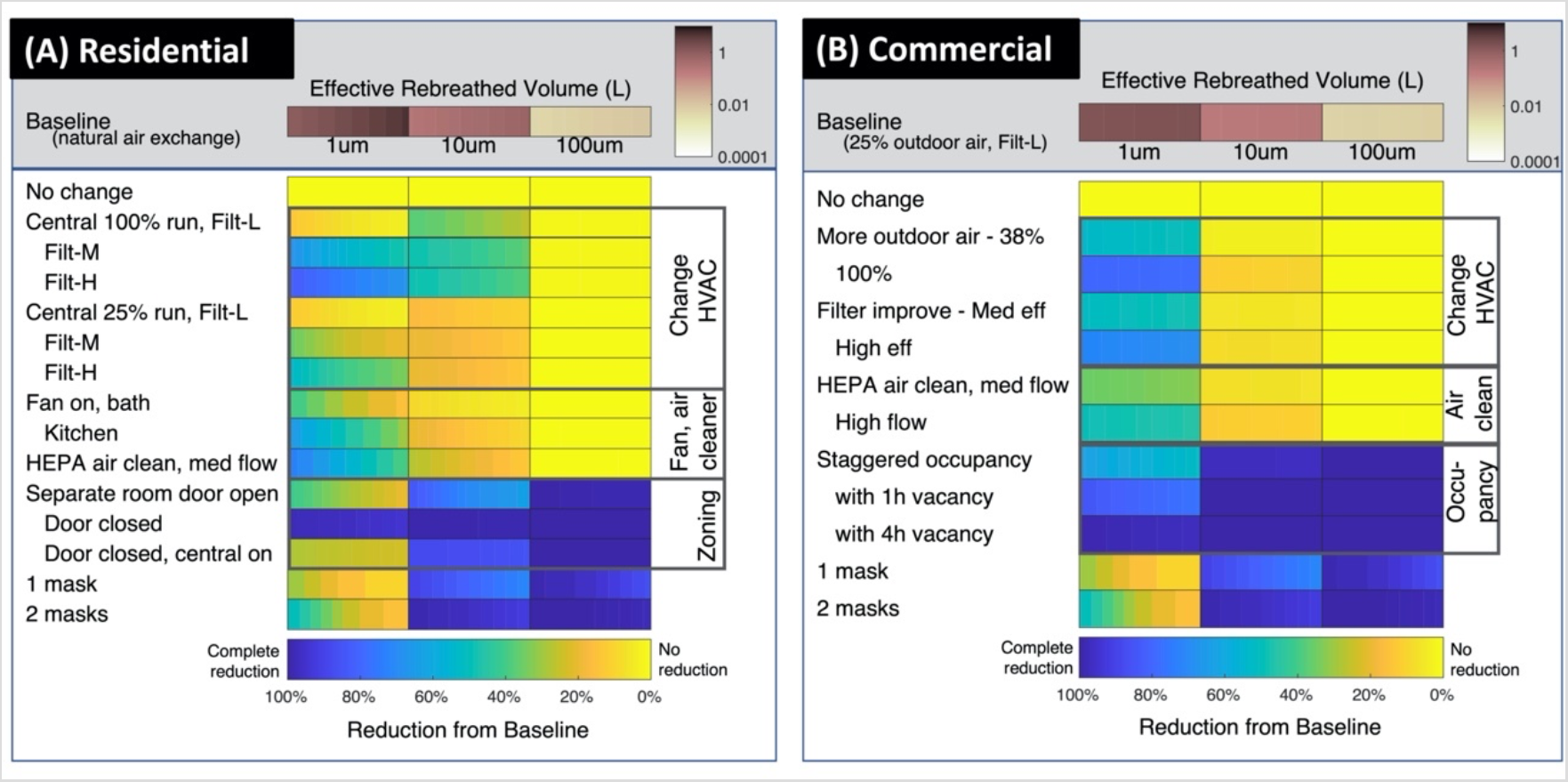
*Person-to-person ERBV for four-hour interactions in (A) residential and (B) commercial settings, along with reductions due to mitigation measures. Horizontal shading in the individual rectangles shows realistic variability in environment and facilities, such as different floor areas (SI). Homes and commercial spaces have similar floor areas in these simulations. In residential settings, “Central” means operating the central air handlers, common in the United States, continuously (100%) or with a 25% runtime. Most of these systems filter air but do not provide fresh air from outdoors; ventilation is provided with bath and kitchen exhaust fans. The situation differs in commercial buildings, where continuously operating central air handlers supply fresh outdoor air as well as filtering recirculated air. “Air cleaner” refers to portable, stand-alone air cleaners with high-efficiency filtration. SI contains additional details, along with 15-min and 1-h figures*.

The upper portion of the figure shows how size-dependent losses affect rebreathing: ERBV_1_ is two orders of magnitude greater than ERBV_100_. This difference does not imply that 1-μm particles have the highest infectivity. If exhaled air contains few or no pathogens of this size, then the efficient transport indicated by high ERBV_1_ is unimportant. The high value does indicate that even a small release of pathogens in m and 10-1-μm particles would be easily transmitted to a recipient. Likewise, the lower value of ERBV_10_ does not indicate unimportance. Particles of 10-μm diameter might be transmitted less efficiently, but this size range could still contain most of the infective particles. The importance of each particle size is different for each pathogen, and is unknown in an emerging situation. We evaluate fractional reductions in ERBV, which does not vary with disease.

The lower part of each figure shows rebreathed volume reductions by ventilation, filtration, and occupancy or zoning measures. The lowest rows show the effect of cloth or surgical face coverings for comparison, with uncertainty in efficiency shown as a range of shading. For mechanical measures (ventilation and filtration), achievable reductions depend on the fraction of time operating, flow rate compared to room volume, and filtration efficiency. Many common filters remove particles of 10 μm diameter and larger, with efficiencies improving at higher filter ratings; lower-rated filters do not remove 1-μm particles.

ERBV_100_ is not noticeably reduced with any ventilation or filtration strategy, because these large particles are lost by deposition more quickly than they can be removed mechanically. Offsetting occupancy, wearing face coverings, and separating occupants between rooms—even with doors open— does reduce ERBV_100_.

ERBV_1_ can be reduced with many ventilation and filtration strategies. Ventilation brings fresh air into the environment, while filtration recirculates and cleans air. Reductions in ERBV_10_ by mechanical measures are intermediate between ERBV_1_ and ERBV_100_. Some effective mitigation measures would be neglected by assuming that 1-μm and 10-μm particles are unresponsive to mechanical means like large, 100-μm droplets. In residences (Fig. 3A), kitchen range hoods reduce ERBV_1_ by 30-40% and bath fans, with about half the flow, by only 15-30%. When central-air units operate continuously with medium- or high-rated filters, reductions are 15-55%; lower operating time decreases those benefits. Separating individuals between rooms gives moderate reductions, while closing doors between them is the best protection as long as central air handlers are not operating. Offices have more closely controlled ventilation than do homes, and a narrower range of ERBV and mitigation effectiveness. Increasing the amount of outdoor air supplied and improving the filter both reduce ERBV_1_, while ERBV_10_ reductions are lower because the baseline already includes some removal of 10-μm particles. Staggered occupancy, in which one person enters after another leaves, reduces ERBV_1_ similar to medium-rated filtration. Vacancy periods increase the reduction.

Deposition loss rates are a key reason that 1-μm and 10-μm particles differ in baseline ERBV, and also explain differences in the ventilation effectiveness shown in Figure 3. Deposition loss rates for 10-μm particles are similar to or greater than air exchange rates, so total removal is less influenced by intentional ventilation changes. In comparison, removal of 1-μm particles is dominated by air exchange and easily altered by ventilation. A good understanding of indoor deposition rates therefore underlies quantification of ventilation effectiveness, but these loss rates are infrequently measured, and measured deposition is usually faster than theoretical predictions (30).

The difference between ERBV_1_, ERBV_10_ and ERBV_100_ offers the possibility to determine particle sizes most likely involved in transmission through retrospective analysis. For example, staggered occupancy (one four-hour shift following another) reduces ERBV_1_ by about 60%, but ERBV_10_ by over 99%. In an emerging disease outbreak, the infectious nature of 1-μm versus 10-µm particles might be elucidated by seeking situations in which an index patient infected others in the same shift, and did or did not infect others in the next shift. A similar epidemiological exploitation has been proposed earlier (31).

## Mitigation in meaningful ranges

Thus far, we have presented ERBV in baseline situations and discussed methods to reduce those values. It is also essential that mitigation measures reduce the risk of infection, not just the dose, because each strategy carries some cost or inconvenience. Ideally, one would be able to calculate the baseline and mitigated doses and evaluate them against a dose-response curve. In the face of a novel pathogen, neither the transmitted dose nor the dose-response curve is known, and only classical disease models can provide some guidance.

Dose-response curves for many viruses have similar features. Human and animal responses typically show a zero risk of infection below the minimum infectious dose, and near-certainty of infection above infectious dose 95% (ID_95_, also called “saturated”). If the baseline situation results in a dose below the minimum infectious value, then mitigation measures are not required. Conversely, if the baseline exceeds ID_95_, mitigation does not effectively reduce risk without a significant reduction in dose. Between the minimum infectious dose and ID_95_, dose-response curves often have a sigmoidal shape. At the point where the dose is likely to infect 50% of susceptible individuals (ID_50_), infection risk rises approximately linearly with the logarithm of dose. Mitigation measures are therefore effective only when applied over this responsive portion of the curve, and efficacy depends on both the baseline risk relative to ID_95_ and the range, or width, of infectious doses spanned by the sloping portion of the infection curve. Neither the baseline risk nor the width of the dose-response curve is known in an emerging situation, and in fact is often unknown for well-studied pathogens.

Figure 4 illustrates risk reductions beginning with four baseline risks, where the reduction in dose (*x*-axis) corresponds to the change in ERBV. A dose-response curve with the shape found for SARS-CoV-1 (32), which is closely related to SARS-CoV-2, is used to estimate remaining risk. A wider dose-response curve, not specific to any pathogen, also appears to illustrate how risk would respond for a disease with different characteristics (Section S.9, Fig. S.5).

The upper portion of Figure 4 shows the reductions possible from the measures in Figure 3. Risk via ERBV_1_ is reduced by many mechanical measures in residential situations, and most measures in commercial situations. Except for occupancy strategies, many measures do not have a large effect on risk via ERBV_10_ in commercial settings. Ventilation strategies do reduce risk via ERBV_10_ in residential settings. When the original risk is very high (95%) and the dose-response curve is wide, the large reductions needed to achieve meaningful reductions are not possible with any mechanical measures.

**Figure 4.**
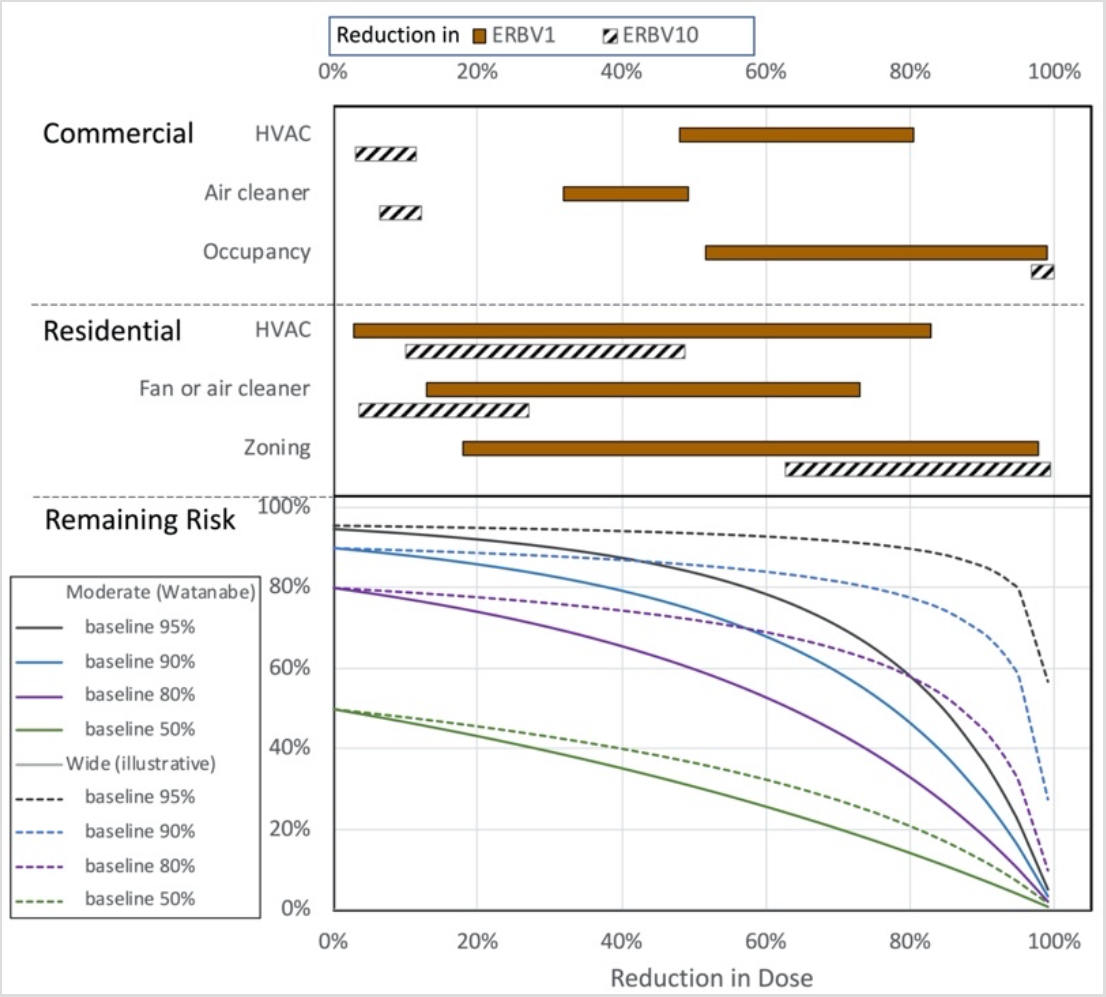
*Possible changes in risk associated with reductions in ERBV. Percentage reductions in ERBV (upper x-axis) are the same as percentage reductions in dose (lower x-axis), and can thus be associated with risk remaining after a specific dose reduction. Baseline risk and dose-response curve width are not known, and uncertainty in risk reduction is demonstrated with different scenarios. Labels on ‘Commercial’ and ‘Residential’ measures correspond to the specific measures grouped in Figure 3, and ranges for each category cover all measures and situations. ERBV_100_ is not shown because most of the mitigation measures have no effect*.

The dose-response relationship is not known in an emerging disease outbreak. Observations of rebreathed volume can serve as a proxy for dose during early decisions about mitigation. When ERBV is comparable to another situation in which infection has spread widely, mitigation measures that give at least order-of-magnitude reduction should be implemented. Identifying ERBV values when infection does and does not occur could suggest the width of the curve, even if uncertainty in ERBV were a factor of three (about 10^0.5^).

## Practical Uses of Effective Rebreathed Volume

By acknowledging that particle size is the cause of differences in transport, ERBV avoids the legacy “droplet” versus “aerosol” dichotomy. We propose that the following steps would have lessened some of the economic impact associated with the COVID-19 pandemic:

1. *Building designers* would have determined ERBV_1_, ERBV_10_, and ERBV_100_ at the time of commissioning, providing values that quantified both normal and transmission-minimizing circumstances.
2. *Epidemiological studies* would immediately exploit known differences in ERBV_1_, ERBV_10_, and ERBV_100_ to identify particle sizes associated with infection as soon as outbreaks emerged. They might also identify ERBV associated with saturation and approximate widths of dose-response curves. Effective interventions could then be better targeted.
3. *Facility managers* could evaluate venues, for example, comparing ERBV for different rooms or for indoor versus outdoor locations. As information emerged, they would be guided in their determination with values of ERBV that were known to be saturated and safe.
4. *Public health messaging* would include ERBV so that each individual could make informed choices about interactions based on relative and overall risk acceptance.

These types of evaluation have all been conducted casually during the COVID-19 pandemic. We developed the size-dependent ERBV metric to provide rigor to such informal evaluations, to isolate the environmental component of the chain of infection, to identify limiting uncertainties like indoor deposition rates, and to provide a framework that supports rapid response in future outbreaks.

## Materials and Methods

Exhaled volume is treated as a conserved tracer with losses that depend on particle size. All equations for transport of a contaminant in fluids can be used to predict the exhaled volume per volume of air. A challenge in developing comparative transport metrics has been the limited literature describing the travel of contaminants within a few meters of a source or emitter, which we call “source-proximate transport.” A contribution of this work is therefore a review of modeling approaches to estimate rebreathed volume. The models chosen are described here, and equations and further justification appear in Supporting Information.

The models ultimately chosen are simple, yet they capture the major factors that affect contaminant transport: distance from emitter, accumulation in confined spaces, dispersion rate, and the influence of other loss rates including mitigation measures. The simulations can therefore be used to compare expected values of rebreathing, even between very different environments such as within and outside of structures. The chosen models also do not rely on specialized inputs such as surface temperatures or roughness, or detailed interior geometries. Such requirements would preclude general recommendations and comparisons among environments.

Most environments are more complex than the simple representations employed here, so contaminant concentrations might vary spatially. However, stochastic variations do not invalidate comparisons between expected values of rebreathed volume, which are averaged across the entire environment. If those variations are not observable or predictable, they cannot be manipulated to reduce risk, either.

For outdoor interactions, we used a steady-state Gaussian plume equation (33) over a range of atmospheric stability conditions. The Gaussian plume is typically not used to describe transport over short distances, because contaminants travel in irregular packets. However, average concentration values do follow the expected shape, even 2 m from the emitter (34, 35), and the distribution of concentration due to sporadic transport on short time scales can be described probabilistically (36). We therefore 4) combined the Gaussian-predicted concentrations with an intermittency enhancement (36) as a worst case. This choice is discussed in more detail in SI S.2.

For indoor interactions, we used a well-mixed zone model, which assumes that concentrations are the same throughout each zone. The model was cast in a matrix form (37) to simulate multiple zones, including connections through central air systems (SI S.3). Removal by filtration was applied at the inlet of the central air system. When natural infiltration dominated air exchange, we accounted for reduced effectiveness of mechanical ventilation (38). Few reports quantify deviations from the well-mixed zone assumption other than proximity effects. Variations in indoor cooking smoke concentrations can be about a factor of two from lowest to highest (39). A simulation of stochastic variations in infection rate resulting from interzonal transport gave about 40% variation (37). No evidence suggests that expected values from well-mixed simulations are biased.

Elevated concentrations occur near emitters indoors. We simulated indoor proximity effects with a point-release model (26), modified to represent a continuous source (27) (SI S.4), with dispersion parameters dependent on air-change rates (27). Concentrations simulated by the point-release model agreed with steady-state, well-mixed values at distances far from the emitter or when dispersion was similar to outdoor values. Figure 2B and Table 1 show worst-case rebreathing rates, when the emitter and recipient breathe at the same level.

We interpreted the few available measurements of the proximity effect in terms of ERBV (SI S.4) and compared them with the point-release simulations, as shown in Figure 2B. The effect of indoor proximity has been simulated with computational fluid dynamic models, but we did not find quantified source or breathing rates that would allow interpretation in terms of rebreathed volume. Computational fluid dynamic studies often examine situations with particular ventilation, furnishing, or occupancy features; they would be useful to a broader understanding of ERBV by including enough variation to allow generalization.

Particle removal by deposition is the reason that ERBV differs among particle sizes, and also a cause for differences in the effectiveness of mechanical mitigation measures. Selection of appropriate deposition rates thus affects all conclusions. We use the theoretical model by Lai and Nazaroff (40) to provide central values. However, measured deposition is often faster than model predictions (41), especially for particles smaller than 1 μm and in occupied houses. Uncertainties in Fig. 2C come from observations (42, 43) and are discussed further in SI Section S.5 and Table S.2.

We identified a range of baseline floor area and air exchange rates for residential and commercial situations (SI S.7, S.8). Air exchange rates include those recommended by the American Society of Heating, Refrigerating and Air-Conditioning Engineers (ASHRAE) (44, 45) as well as measurements. For each baseline case, each mitigation measure was applied and then the reduction in ERBV was calculated to obtain the range of reduction percentages in Figure 3. Filter efficiencies are taken from ASHRAE Standard 52.2–2017 (46) with ratings as described in Supporting Information.

## Data Availability

All input data and equations are given in supporting information.

## Acknowledgements

Mj Riches created the artistic rendering of Figure 1. Helpful comments on the manuscript and its development came from David Dandy, Greg Hickey, Joshua Keller, Marilee Long, Steven Simske, and Ander Wilson. The author team acknowledges multidisciplinary interactions supported at Colorado State University that fostered collaborative problem-solving.

## Contributions

Conceptualization (TCB), Methodology (TCB, ABL, DKF, PWF, JRP, JMH), Validation (SHJ), Cross-disciplinary integration (KMF, SV), Writing – Original Draft (TCB, ABL, DKF, PWF); Writing – Review and editing (All).

## Data and materials availability

All input data and equations are given in supporting information.

## Supporting Information

### Supplementary Text

#### S.1 RBV calculations

Methods of calculating rebreathed volume (RBV) are described first (Sections S.1-S.4). Then, modifications that account for deposition losses are discussed (Section S.5).

Exhaled volume is treated as a conserved tracer (*Bex* in the equations that follow). All equations for transport of a contaminant in fluids can be used to predict the exhaled volume per volume of air (*b*_ex_) as a function of location and time: *b*_ex_(*t, x*). Although exhaled volume can be modeled in three dimensions, only a single spatial coordinate is used in the following discussion for simplicity.

The recipient breathes at a rate *p*, and the volume of formerly exhaled air that is inhaled by the recipient during an infinitesimal time *dt* is *p b*_ex_ (*t*,*x*)*dt*. Total rebreathed volume during residence time Δt is thus

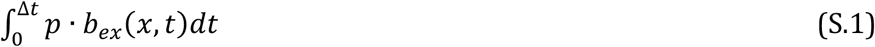

Units of volume per volume for *b*_ex_ can be difficult to conceptualize and the dimensionless quantity *b*_ex_ might allow human errors during the creation of a new model. One alternative is modeling a mass concentration, which has a fixed value *Cex*(kg/m^3^) in the emitter’s exhaled air. The inhaled total mass can then be divided by *C_ex_* to obtain the rebreathed volume.

We use a value of 8 L min^−1^ for the human breathing rate, *p*. This value is common throughout exposure literature (1). This value is greater than the “minute ventilation” used in the medical field (typically 6 L min^−1^), which is a resting rate that does not include activity.

Some studies report steady-state (unchanging) concentration values (*c_ss_(x)*) that result from a constant emission rate, or source, *S*. The ratio *c_ss_(x*)/*S* at any location is a transport function with units of s m^−3^, which quantifies the response of concentration to a unit increase in source strength, assuming that loss processes are linear in concentration. Rebreathed volume during time Δt is

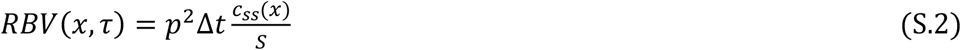

where one of the *p* values accounts for exhalation by the emitter (the source) and one for inhalation by the recipient. The ratio *c_ss_*/*S* allows the use of measured, modeled, and analytically predicted concentrations. Concentrations could also be simulated with computational fluid dynamic models, as suggested by *Sze To and Chao*(2). When concentration is changing rather than steady, the calculation of RBV requires an integral formulation.

#### S.2 RBV outdoors, from plume equations

For outdoor plumes, concentration values at any distance downwind follow a normal probability distribution, with the peak directly downwind from the emitter. Dispersion is described by plume widths that increase with distance, depend on atmospheric stability, and are fit to time-averaged, measured data (3, 4). The standard steady-state Gaussian plume equation is (3):

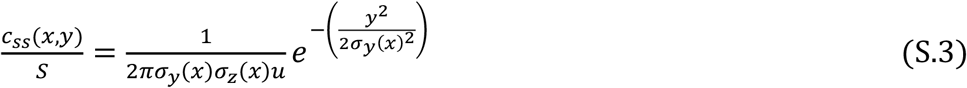

In this equation, *x* is the downwind distance of the recipient, and the dependence on downwind distance is expressed by σ_y_(*x*) and σ_z_(*x*), which can be thought of as is the crosswind distance, and emitter and recipient are assumed to be at the same height for the most conservative case. When the recipient is directly downwind, the exponential term becomes unity.

Literature on parameterizing σ_y_ and σ_z_ under different atmospheric conditions is extensive. In stable conditions, there is less dispersion, σ-values are smaller, and concentration is greater. We use the σ-values recommended by Briggs, fit to curves known as the Pasquill-Gifford method, as tabulated by Hanna (3). Plume concentrations were calculated for each distance over the entire stability range (classes A through F) for urban conditions and for most open-country conditions. “Open-country” dispersion differs because surface obstacles are not as high and turbulence is relatively lower. We excluded open-country slightly- and moderately-stable conditions (classes E and F), reasoning that the emitter and recipient themselves would enhance the turbulence. We also explored other dispersion relationships (5) as a function of boundary-layer height and vertical velocity; these gave rebreathed volume values in the same order of magnitude.

There is a concern about using Gaussian plume models to describe transport over distances of only a few meters. Values of σ_y_ and σ_z_ are generally provided for distances of 100–10,000 meters downwind of the emission source. The term “near-source,” in dispersion literature, is associated with distances of 50–100 m. We found very little support for estimating what we have come to call “source-proximate” dispersion: transport occurring over distances of less than 10 m, relevant to person-to-person transport and to exposures of individuals very near a hazardous material release. Contaminants travel in irregular packets, so the normal or Gaussian shape is found only after averaging (6, 7), typically over several minutes; the Gaussian plume is usually applied more than 50 m from emitters. However, average concentration values do follow a standard plume shape (8, 9), and the concentrations resulting from sporadic transport can be described probabilistically (10). We found only one data set of source-proximate concentrations measured by Jones, at 2, 5 and 15 m downwind (7). Average values of *c*_ss_/*S* at 2 m were 0.26 s m^−3^, or 0.65 s m^−3^ when adjusted for electrostatic effects that affect those particular measurements. These values lie in the range for plumes that are neutral or slightly unstable, extrapolating the Briggs fits for σ back to 2 m. Furthermore, dispersion principles are the basis for modeling plumes as a series of filaments for odor transmission in the source-proximate range (9), and those simulation results compare well with the Jones measurements. We conclude that standard plume equations can be used to estimate source-proximate concentrations, with caution. The proximity curves in Figure 2 use the classical plume equations for a receptor directly downwind.

The Gaussian shape of the plume cross-section emerges only after averaging over several minutes. The recipient might intercept whiffs, or plume tendrils higher than the average concentration, over short time scales. We estimated an “intermittency factor:” an increase in the average concentration if the recipient were to encounter *only* the whiffs. Based on the data of Murlis and Jones (10), this increase is about a factor of five, as the transported material is observed only about 15-20% of the time. This factor is quite conservative as these observations are made with time resolution of less than 1 s. The intermittency factor would decrease rapidly with averaging time and disappear after about 20 minutes (11).

For wind speed, 2.5 m s^−1^ (5.6 mi h^−1^) at 2 m height is roughly 3.3 m s^−1^ (7.5 mi h^−1^) is similar to the annual-mean 10 m wind speed in many cities (https://www.currentresults.com/Weather/US/wind-speed-city-annual.php). When all other factors are equal, outdoor ERBV is inversely proportional to wind speed, so the estimates in Fig 2 may be scaled to different wind speeds using (2.5 m s^−1^), where *u* is the wind speed in m s^−1^.

#### S.3 Well-mixed indoor environments

A balance equation for the total exhaled air which is introduced by a human breathing, mixed instantly into a box volume V, and carried out by air exiting the volume at a rate *Q*, is given by:

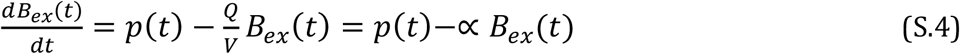

In indoor air quality literature, the ratio *Q/V* is known as an air change rate, with units of inverse time, and is often given the symbol α, as shown in the rightmost representation. Other first-order loss rates, such as deposition (Section S.5), can be added into the final term in the equation.

The relationship for exhaled volume per volume of air (*b_ex_*) can be expressed by dividing by the room volume, V. Balance equations, and their use in modeling indoor environments, have been discussed extensively in the literature, and analytic solutions are not repeated here. Of particular note are solutions for accumulating breath (12), for differing occupancy time of emitter and recipient (1) and for accounting for filtration and personal protective equipment (13, 14).

When multiple, well-mixed zones are connected, the equation for *B_ex_* in each zone *i* is(15):

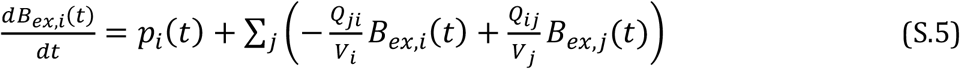

where *Q_ij_* is the flow from zone *j* to zone *i*. In matrix form, each element of vectors 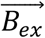 and 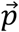 corresponds to a zone:

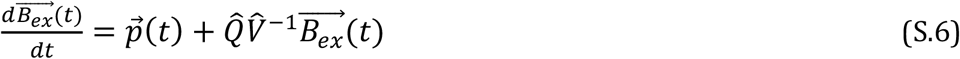

The square flow matrix 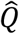 consists of off-diagonal, positive flow elements *Q_ij_* from zone *j* (columns) into zone *i*(rows). The diagonal elements *Q_i,i_* are negative, representing the total flow out of zone *i*. Off-diagonal elements in each row or column must sum to the diagonal element, assuming incompressible flow. The volume matrix 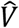 is diagonal only and contains the volumes of each zone. One zone can have a zero concentration and infinite volume to represent the outdoors.

The matrix formulation was coded in MatLab for this work. Other multi-zone models exist, notably CONTAM (16); our representation relies on specifying interzonal flows rather than changing them due to pressures. and enabled the setup of several similar situations with the same mitigation measures. Central air handlers were represented as separate zones to simulate multiple inlets and outlets to and from the same conditioning system. Filtration was represented as a fractional removal at the inlet to the air handler.

The formulation specifies flows between zones or between indoors and outdoors. An inherent assumption is that flows added to each zone do not affect pressures enough to alter other interzonal flows. This assumption is safe when the added supply and return flows are balanced, or when all fresh air is mechanically driven, as is the case in many commercial situations. The assumption is not true in many residences, where the baseline airflow between indoors and outdoors is caused by pressure gradients driven by natural temperature differences and wind. When mechanical flow *Q_mech_* is imposed under these conditions, the added flow *Q_add_* also depends on the natural ventilation rate *Q_nat_* (17):

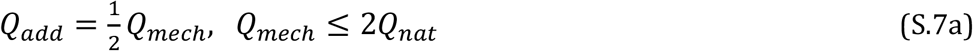

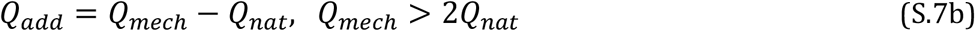

#### S.4 Indoor point-release model

Releases in indoor environments do not immediately mix into the room, so both proximity and confinement effects exist. We simulated indoor dispersion and proximity effects with a point-release model as described in Materials and Methods; additional detail and justification is provided here. The point-release model accounts for the influence of walls using an infinite series of mirror-image sources; this series was calculated until it converged. The use of image sources probably overestimates the retention in air of particles that deposit to surfaces, but we used the model mainly to estimate proximity effects near the emitter, where most of the concentration is due to the original plume and not the reflected plumes.

We simulated a range of room sizes, air change rates, and emitter locations for 1um, 10um, and 100 µm particles, as summarized in Table S.1 (27 simulations for each particle size). Dispersion parameters depended on ventilation rates as derived from measurements by Cheng *et al*. in still rooms (18). The proximity effect was greatest when dispersion was slowest, when the emitter was near a wall, and when the room was small. In real situations, dispersion is probably more rapid than the still-room situations shown here, so the proximity effect would be lower. Ranges of rebreathing from all the simulations are summarized and compared with simple indoor and outdoor models in Figure 2 (1 µm) and S1 (10 µm and 100 µm). The rebreathing rates shown are taken at the same height as the emitter, and thus represent the worst case at each distance.

To illustrate the interaction between proximity and confinement further, Figure S.2 shows the modeled rate of rebreathing as a function of residence time and distance from the emitter for a single room size (6m × 6m) and ventilation rate (0.3 ac h^−1^). The gradient from left to right at less than 1 m distance shows the proximity effect; the gradient from top to bottom at 3 m distance shows the confinement effect; and the contours show the presence of both proximity and confinement effects near the emitter.

The simple point-release model does not capture all air currents and, hence, concentration variations caused by human figures, ventilation flows, and furnishings. Computational fluid dynamic models are more suited to simulate those details. We use simplified models to cover a range of conditions, to increase the generality of conclusions.

We compare our range of breathing rates with those of two studies designed to estimate the proximity effect indoors. Neither of those studies gave RBV, and our interpretation from their results is described here. Licina *et al*. (19) measured intake fraction for particle releases at different locations around a thermal manikin in a test chamber. The release occurred over 10 minutes, after which the source was turned off, so only the 10-minute concentrations are comparable to the point-release simulations. This study provided a total intake fraction for all 10 minutes. To translate this value to an instantaneous rebreathing rate for Figure 2B, we used our well-mixed room model to determine the ratio between ERBV_1_ for time 9-10 min, and ERBV_1_ for time 0-10 min, and applied this to the published 10-minute intake fraction. We used only those measurements with the release 1 m from the manikin, at different heights, but not at the floor. See Section S.10.4 for relationship between ERBV and intake fraction.

Liu *et al*. explored indoor proximity effects using measurements of NO_2_. They defined an exposure index as the ratio between the concentrations at a particular point and that averaged throughout the room. We used our well-mixed model to determine steady-state RBV per minute, and multiplied it by the maximum exposure index reported at each distance from the emitter (Fig. 5 in *Liu et al*.). Like the results of the point-source model, the data points representing the Liu study in Fig. 2 are the worst case. Liu *et al*. (20) also used computational fluid dynamic simulations to explore the transport of different particle sizes, but did not provide enough information to calculate ERBV. We encourage future studies to provide quantitative values of source strengths and concentrations, from which ERBV can be interpreted.

#### S.5 Size-dependent deposition losses

RBV differs from ERBV by accounting for size-dependent losses due to particle deposition and removal strategies. In this section, we summarize deposition loss rates that modify the outdoor and indoor values of RBV.

Particle deposition, or attachment of particles to surfaces, combines several processes that depend on particle size:

1. gravitational settling, or direct interaction between a particle and surface due to gravitational forces
2. impaction, or direct interaction between a particle and surface driven by turbulent air flows that enable the particle to contact the surface
3. interception, or indirect interactions in which turbulent air flows bring a particle close to a surface, enabling particle removal, and
4. Brownian diffusion, in which particle interactions with a surface are driven by random motion of the particle in air.

The magnitude of each loss rate depends on particle size. Particles below 0.1 μm in diameter are dominated by Brownian diffusion losses, while the loss of particles above 10- μm diameter is dominated by gravitational settling. The total deposition loss rate is a combination of these factors, and is often described by a parameter known as deposition velocity v*d*:

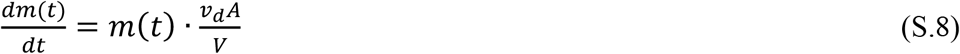

where *m* is the mass of particles of a certain size, A is the area to which they deposit, and V is the volume from which they are lost. The quantity (*V/A*) is the effective height (*heff*) of the volume; the ratio *v_d_ / h_eff_* is a loss rate that has units of inverse time; and the product (*v_d_A*) can be treated as a volume flow exiting the space. These terms are included in the mass balance equations.

Table S.2 summarizes deposition velocities used in calculation. Outdoor values of *v_d_* are the gravitational settling velocity, assuming a density of 1 g cm^−3^ for particles that are mostly water. The fraction lost is estimated in the plume model as:

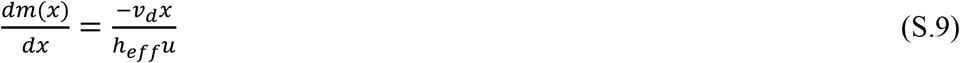

We assumed an effective height of 1.5 m, the approximate mouth height. Deposition losses over source-proximate distances are small; even for 10-um particles, less than 0.2% of particles are lost.

Deposition velocities in the indoor environment have been parameterized theoretically (21), modeled with computational fluid dynamics (22) and measured (23, 24). The theoretical model by Lai and Nazaroff (21) is well-established in the indoor air literature, and its predictions are used as central values. Measured deposition is often faster (i.e. a larger deposition velocity) than model predictions, especially for particles smaller than 1 μm and in occupied houses (25). The assumption of deposition to upward-facing surfaces only is likely acceptable for 10-µm and greater particles, but not for 1-µm and smaller particles.

Table S.2 summarizes uncertainties in deposition velocity based on observations (26, 27) and measurements from the HOMEChem experiment. Computational fluid dynamic models have simulated lower values (22), which are not included in the uncertainty ranges because they do not improve agreement with measured values.

Indoor deposition velocities are similar for particles of 0.1µm and 1µm. Although the 0.1-μm particles settle more slowly than 1-μm particles, they have greater losses by Brownian motion. For this reason, we do not report 0.1-μm particles separately in the mitigation measures; the calculation of rebreathed volume would be nearly identical. Unlike outdoor deposition losses, indoor losses over the period of confinement are significant, equal to or greater than typical ventilation rates, as shown in Table S.2. For 10-μ the 0.1-μm particles settle more slowly than 1-µm particles, they have greater losses by Brownian motion. For this reason, we do not report 0.1-μm particles, these losses are particularly important in determining indoor concentrations. The effect of deposition uncertainties on indoor rebreathing rates is shown in Fig 2C in the main text and further discussed in Section S.11.

#### S.6 Particle removal by face coverings

Reduction of ERBV by face coverings (masks) is included in Figure 3 for comparison with other mitigation measures. Mask efficiency ranges are estimates for the wide variety of face coverings in public use. Low-cost mask testing is becoming widespread (28) but counts particles rather than assessing removal of different particle sizes. Official mask-testing protocols recommend measurement of particles at 0.3 μm. For 1-μm particles, we used the tabulated range of efficiencies from a laboratory that maintained an active and on-line database of tests at 0.3μm size, covering many common materials (29). In our figures, the range of particle penetration for a single mask was 70–95%. Impaction and other mechanisms should stop many 10-µm particles, but they can escape by leaking around the material of an unsealed mask. Oberg and Brosseau (30) reported surgical mask penetration of less 1% to 67% for 3.1-μm particles, with most of the penetration values below than 1%. For 10-μm particles, we used penetration values of 1–30% to account for cloth masks and poor fits. We found no data for 100-μm particles and used penetration of 1%-10%; the highest value accounts for possible breakup of very large particles through the mask. This range probably overestimates penetration but does not affect any of the comparisons.

#### S.7 Residential situations

Inputs for residential simulations are summarized in Table S.3. In the baseline residential case, we assume that the central air handler does not operate, corresponding to mild weather conditions. We also assume that bathroom and kitchen fans are not used continuously.

The floor area covers the most common housing sizes in the United States, representing approximately two-thirds of the U.S. housing stock (31). The ceiling height is a typical height for U.S. residences. If a portion of the home were closed off, the smaller end of floor area and volume, associated with higher ERBV would be appropriate. However, people who might be concerned with infecting each other are most likely to interact in rooms whose connectivity comprises a large fraction of the home, such as a connected living room, kitchen, and dining room.

Recommended ventilation depends on the number of occupants and the floor area of the home. In the most recent guidance, the lowest air flow normalized by volume corresponds to an air-change rate of about 0.3 ac h^−1^ (32), and we use this as a central value.

Air change rates vary widely throughout the housing stock. Characterization of leakiness is often reported as “ACH50”, or flow at 50 Pascals of depressurization by a blower door fan normalized by the volume of the conditioned space. The pressure under testing conditions is much greater than naturally-occurring pressure differences; the measured flow must therefore be adjusted to natural conditions. This adjustment depends on factors such as climate and house height, but a common rule of thumb is dividing ACH50 by 20 to obtain air-change rates under normal conditions (33). A database for the United States (34, 35) found that 3% of homes had ACH50 below 2, corresponding to a natural air-change rate of about 0.1 ac h^−1^, and we use this value as the lower end of the range. Average ACH50 for homes participating in weatherization programs, and hence older, have been reported as around 20 (36), corresponding to about 1 ac h^−1^ under natural conditions, and we use this value as the upper end of the range. The range (0.1–1 ac h^−1^) spans about a factor of three below and above the ASHRAE 2019 target. Values above 1 ac h^−1^ in normally-ventilated spaces are not supported by observations in the United States. Estimating air-change rates with tabulations of ACH50, without adjustment, would greatly overestimate ventilation.

Table S.4 summarizes mitigation measures for residential cases and provides justification. The “Label” column and order corresponds to the labeling in Figure 3. The “Description” column gives fan or air handler flows and filter ratings. Filter ratings are reported according to the Minimum Efficiency Reporting Value (MERV) from ASHRAE Standard 52.2–2017 (37). We used central values of particle removal for each rating. MERV ratings cover three particle sizes: 0.3–1.0 μm, 1.0-3.0 μm, and 3.0-10 μm. The 1.0-3.0 μm range is more representative of removal efficiency at μm (38), and we use that range for 1-μm particles.

#### S.8 Commercial situations

Inputs for the commercial baseline simulations are summarized in Table S.5. Central air handlers often run continuously to provide fresh air, unlike in residential situations. The floor area is an open office arrangement, chosen to be similar in size to the residential situation. While there is no requirement in the United States for square footage per person, several real estate websites indicate that open office areas should provide about 100 ft^2^ (10 m^2^) per person, so this area would house about 20 people. Conditioning systems for this floor area would be sized at about “4 tons” or about 14 kW, and the total airflow for this size would be about 1600 cfm.

A common conditioning and ventilating approach for commercial buildings is to recirculate most of the building air through a system located on the roof (“rooftop unit”), and to introduce fresh air from outdoors (“outdoor air”) into this rooftop unit. Thus, more air is supplied to the conditioned space than is withdrawn from it, and the flow is balanced when building air exits through cracks. The baseline is the minimum required by ASHRAE Standard 62.1–2019 (39). The filter rating of MERV 8 is also the minimum required by ASHRAE 62.1–2019. Because the baseline commercial case has some outdoor ventilation air and some filtration, mitigation strategies that appear identical have different reductions than they do in residential situations.

Table S.6 summarizes mitigation measures for commercial cases, with the “Label” column corresponding to the labeling in Figure 3. The “Description” column provides fan or air handler flows and filter ratings.

#### S9. Influence of baseline dose on effectiveness of mitigation measures

Decreasing ERBV and hence dose of infectious particles cannot alter risk below the minimum effective dose. Above the dose where 95% or more of individuals are infected (ID_95_), moderate reductions do not decrease risk either. Further, risk of infection approximately responds to the logarithm of dose between those points. We use two illustrative dose-response curves to demonstrate how the starting point (baseline dose and risk) and the slope of the dose-response curve affect expected risk reductions.

We use the summary of Watanabe *et al*.(40), who synthesized several studies on SARS-CoV-1, to provide illustrative dose-response curves. SARS-CoV-2 is similar to SARS-CoV-1, but we do not assume that the two viruses have the same dose-response curve. We use only features of the curve’s shape to explore how mitigation measures might reduce risk. Watanabe *et al*.(40) explored both exponential and beta-Poisson distributions to fit observed risk of infection. They recommended the exponential distribution because the latter, more complex distribution did not have a statistically better fit to the data. However, the exponential distribution has a fixed width in log-space, so for this exploration we used the beta-Poisson distribution.

We created two distributions with identical values of ID_50_, but different widths (Fig. S.5). The narrower curve (“moderate” in Fig. 4) has a factor of 100 between the doses associated with 5% risk and with 95% risk, informally called “2 logs.” This factor is similar to that of the exponential curve in Watanabe. In the wider curve (“wide, illustrative” in Fig. 4), the distance between 5% risk and 95% risk is a factor of 3100 (“3.5 logs”), similar to curves estimated by Kitajima *et al*. (41) for H5N1.

#### S.10. Chain of infection, rebreathed volume, and the Wells-Riley equation

The Wells-Riley relationship (42, 43) combines characteristics of a population, infectors, and the environment into a mean number of infections μ:

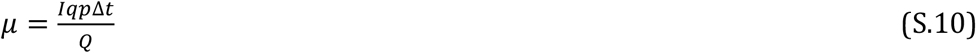

where *I* is the number of infectious disease carriers in a space, q is the emission rate of infection quanta, p is the human breathing rate (average 8 L min^−1^), Δt is the time of the recipient’s exposure, and Q is the ventilation rate. This value of μ is then used (S.11) in the Poisson distribution to determine a probability of infection.

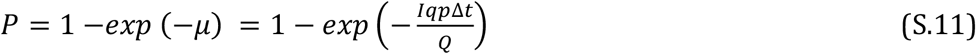

The Wells-Riley equation has often been used in situations where the number of carriers and the ventilation rate is known or can be guessed. It has also been related to the reproductive number *R0* and the susceptible-infectious-recovered (SIR) structure (44). Here, we suggest a slight reformulation of the mean infection rate to distinguish separate roles within the chain of transmission: reservoir (carriers), environment, and host (recipients).

For a single emitter, the concentration of infectious material in a ventilated room at steady-state is *q/Q*, and the dose is the inhaled volume times the concentration: (*q p t/Q*). The emission rate of infection quanta is a characteristic of an individual carrier and is an unknown in an emerging situation. Quantifying a carrier’s emission rate as Θ_c_, quanta emitted per volume exhaled,

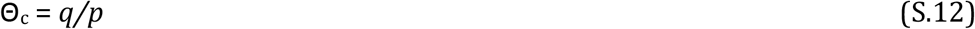

the mean number of infections considering all carriers can be written:

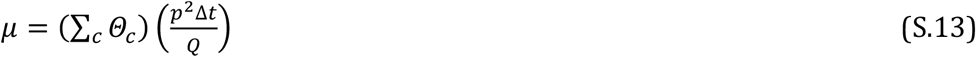

where Σ_c_ indicates a sum over all carriers. When the room is in steady state, meaning that no concentrations change, the term (*p^2^ Δt*/*Q*) is the rebreathed volume (RBV). Even in a more complex ventilation situation, the dose obtained from a single carrier is *Θ_c_* multiplied by the rebreathed volume.

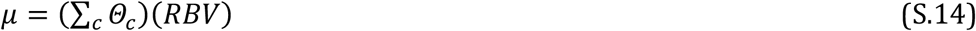

The influence of particle size will be discussed later. Until then, we acknowledge that particle size does have an influence by using ERBV instead of RBV in each equation.

Next, we add some terms that account for other variations. Some infectious individuals might exhale more or less volume, and likewise some susceptible individuals might inhale more or less. These factors can be represented by adjustment ratios between the individual’s breathing rate and the average human breathing rate: γ_c_ for the carrier, γ_s_ for the susceptible host, so that

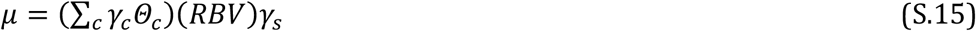

A major insight from Wells’ work was the use of an infection quantum. This concept liberated the analyst from quantifying exactly how many virions or other pathogens were emitted, or how many were required in a minimum dose. The only requirement was determining the emission rate of just enough pathogens to cause an illness, which could be retrospectively calculated from epidemiological observations. However, this use of indivisible quanta implies that the minimal infective dose is identical for every recipient. The infectious dose of SARS-CoV-2, or any emerging disease, for humans is unknown and dependent on numerous host factors, including receptor binding and distribution. It is possible that some hosts are susceptible to a lower dose of the virus than others. Again using a ratio *φ* between the minimal infective dose for the susceptible individual and that for an ‘average’ individual,

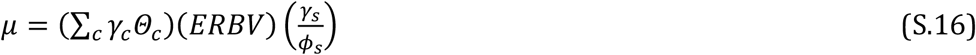

The mean infection rate is now separated into three distinct components that parallel the divisions in Figure 1, as summarized in Table S.7.

The purpose of including the ratios γ_i_, γ_s_ and *φs* is not to call for more information that is difficult to acquire, particularly for a novel disease. When characteristics of individual emitters are not known, equation (S.14) can be used. However, Equation (S.16) makes explicit the roles of carrier characteristics and individual susceptibility when selecting situations for interaction. For example, if breathing rates of both emitters and recipients increase by about a factor of 7 in exercise rooms (γ_i_ = 7, γ_s_ = 7), then achieving the mean infection rate found in a room of sedentary people would require decreasing person-to-person ERBV by a factor of 49, even without accounting for changes in exhaled infection quanta. Likewise, if it is found that individuals between ages 70–75 are infected at one-quarter the dose of the average population, ERBV for their situations should decrease by a factor of 4 compared with another situation that has an acceptable mean infection rate for the average population.

##### S.10.1. Particle size dependence

The quantity *ERBV* replaced the simpler *RBV* to communicate the influence of particle size on pathogen transport and loss. Other parameters in Equation (S5) or (S8) also depend on particle size, especially Θ*c*. Emission rates differ with size, as do the location of deposition within the susceptible host, the susceptibility, and hence the minimal infective dose. Using the subscript *d* to indicate particle diameter, the simpler and more complex equations become, respectively,

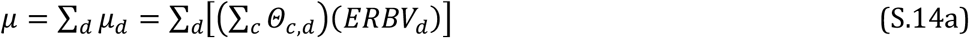

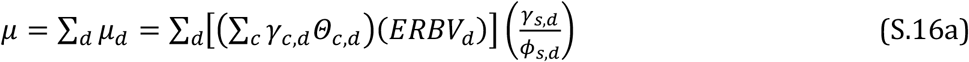

This size dependence may also seem to impose a need for information that might be scarce. The text shows, however, that ERBV_d_ can vary by one or two orders of magnitude between sizes. This difference in size-dependent contributions to *μ* does add complexity, but it also isolates a variation that might be exploitable in analysis.

##### S.10.2 Probability of infection, mean infection rate, and accumulated interactions

The Wells-Riley equation was originally developed to study a school measles outbreak – a situation where exposure occurred in a well-defined, constant situation. A single mean infection rate was appropriate for that situation. When carriers may be encountered through a range of situations, such as a series of university classes or a shopping trip, a summation of infectious contributions through each interaction (subscript *j*) may be more appropriate.

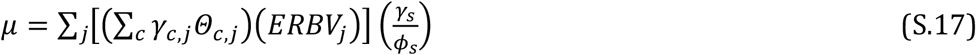

The characteristics of the recipient remain separate, as do the terms for carriers and transport in *each* environment. Only after combining the accumulated interactions would it become appropriate to include the mean infection rate in the Poisson probability equation (S.11). Interactions that dominate the sum are most likely to be implicated in susceptibility. Determination of the time period over which doses might accumulate and collectively contribute to infection also requires investigation.

##### S.10.3 RBV versus rebreathed fraction

The rebreathed fraction can be calculated from mass-balance equations, as well as estimated from CO_2_ concentrations, even in complex ventilation situations (12). Rebreathed fraction is the closest published measure to RBV. Rebreathed fraction differs because it includes the exhalations of all breathing individuals in the building instead of just one. RBV also differs because it is not a fraction of exhalation, but the total exhaled and rebreathed volume, which is proportional to overall dose. RBV could be estimated from CO_2_ concentrations measured in real situations if there were only a single individual in the building, or if every breathing individual were assumed to contribute equally.

##### S.10.4 RBV versus intake fraction

Intake fraction is the amount of emitted mass that is subsequently inhaled by individuals. RBV is proportional to individual intake fraction (iF), as defined by Nazaroff (1). Since the emission is *p* (the human breathing rate) multiplied by the time over which the emitter is active (Δt_emit_), RBV is just

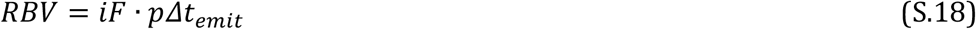

The main difference is that RBV is not a fraction of exhaled air but a total dose.

#### S.11. Influence of deposition loss on effectiveness of mitigation measures

When deposition loss rates are significant, they can dominate total removal, and increasing ventilation may have a small incremental effect. This is not a new finding (e.g. (45)), but is so important to the ability to mitigate ERBV_10_ that it is expanded here. When ventilation is added to a room, the fractional increase in ventilation flow (*δQ/Q*) produces a fractional decrease in steady-state concentration (*δC/C*), if ventilation is the only removal. This change in concentration is the expected reduction. For contaminants that have other removal mechanisms, adding ventilation flow produces a lower reduction in concentration. We define ventilation efficiency as:

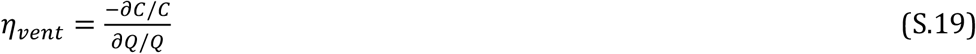

or the ratio of the actual change in contaminant concentration (or ERBV) to the change that would be expected without deposition. Ventilation efficiency depends on the original ventilation rate in addition to deposition and other losses, and is graphed in Fig. S.6 for a range of deposition velocities and three baseline ventilation rates. The range of deposition velocities in Table S.2 is also shown on the figure. For 10-µm particles, ventilation efficiency ranges from 1–25%.

## Figures

**Figure S1.**
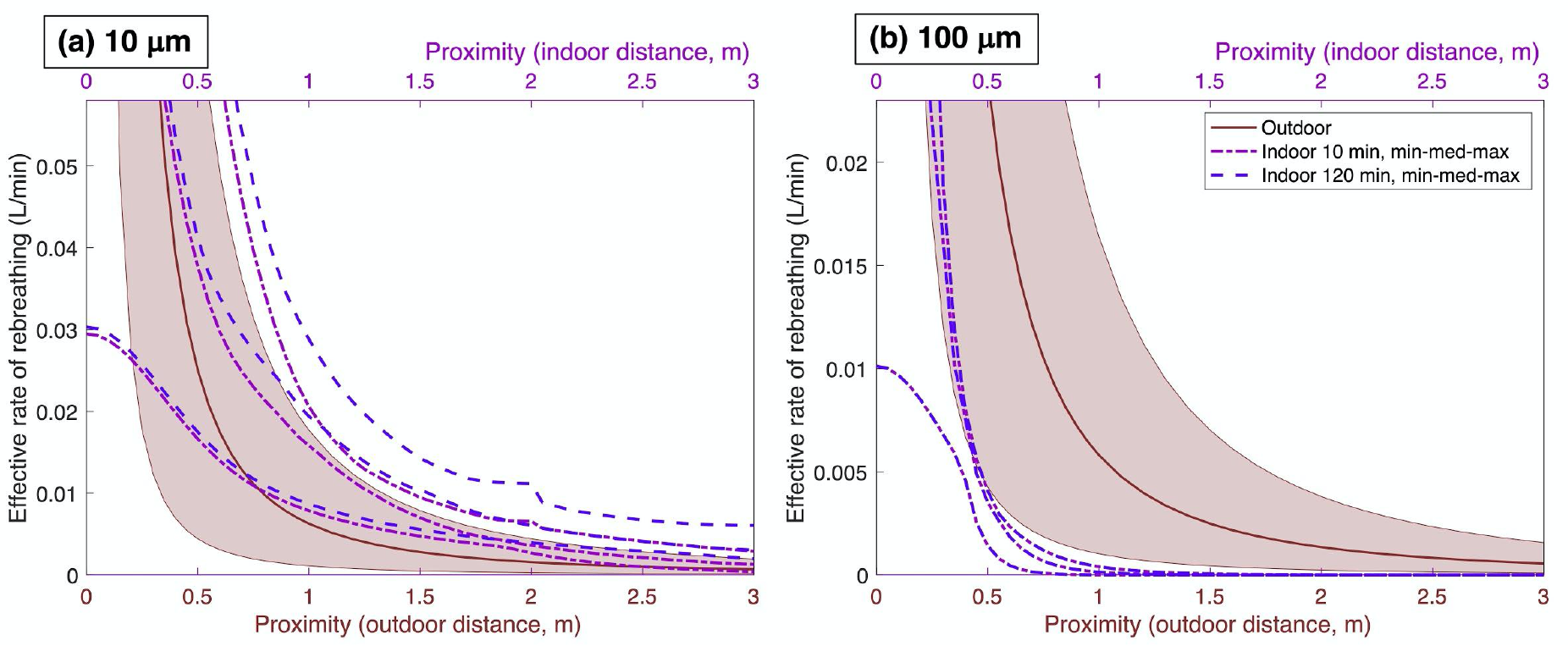
Comparison between indoor and outdoor proximity effects. Figure is for (a) 10-µm and (b) 100- µm particles (comparable to Figure 2(b) in text). 100- µm particles travel farther outdoors because they are carried by wind. For 10- µm particles, highest ERBV occurs in small rooms, which disappear from the summary curve when the maximum possible distance from emitter is reached, creating the discontinuity shown in the dashed lines.

**Figure S.2.**
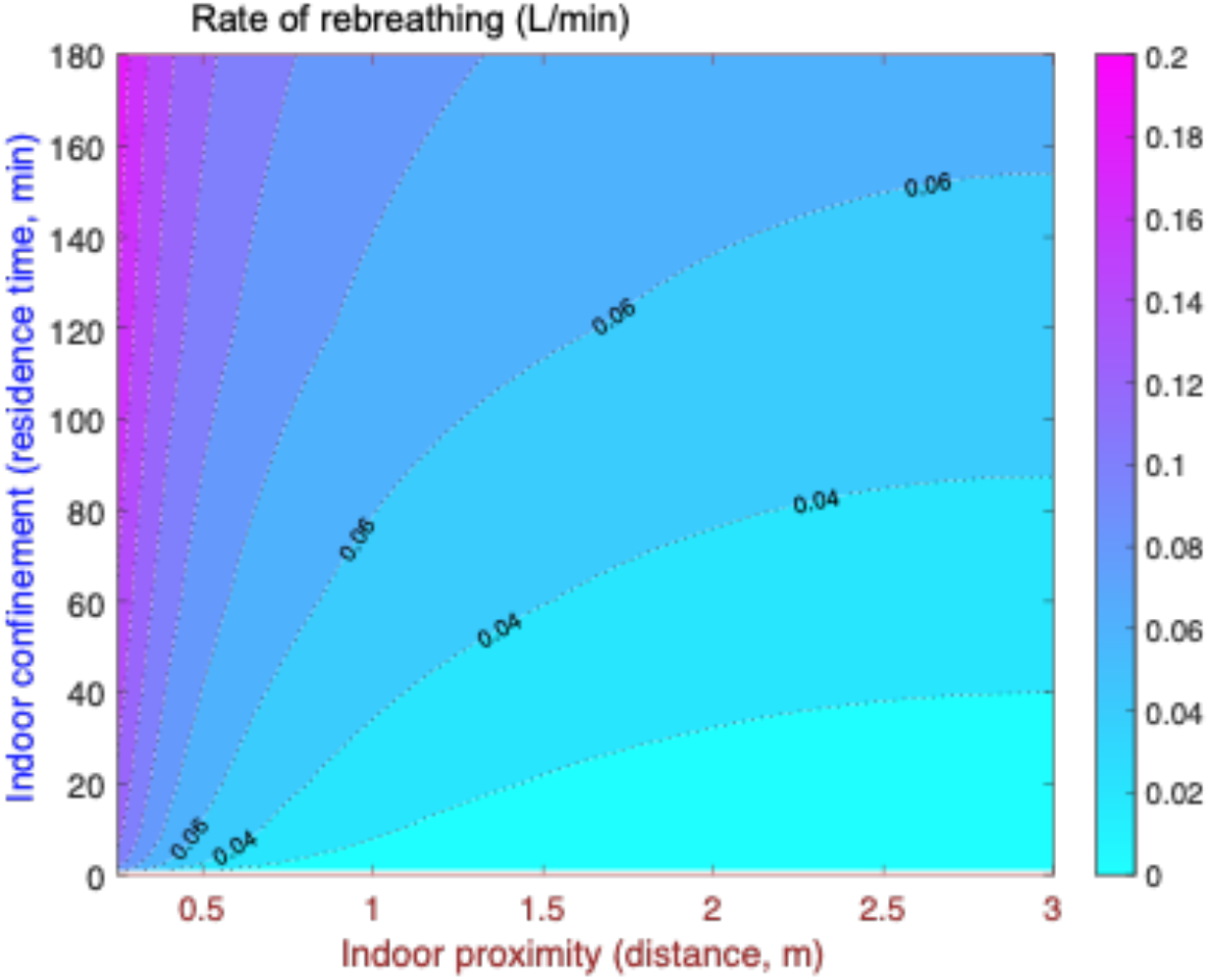
Rebreathing rates simulated by the indoor point-release model. Figure is for 1-µm particles, 0.3 air changes per hour, and 6mx6m room with emitter in the center. The marked contours bound the rate of rebreathing in a well-mixed case at steady-state.

**Figure S.3.**
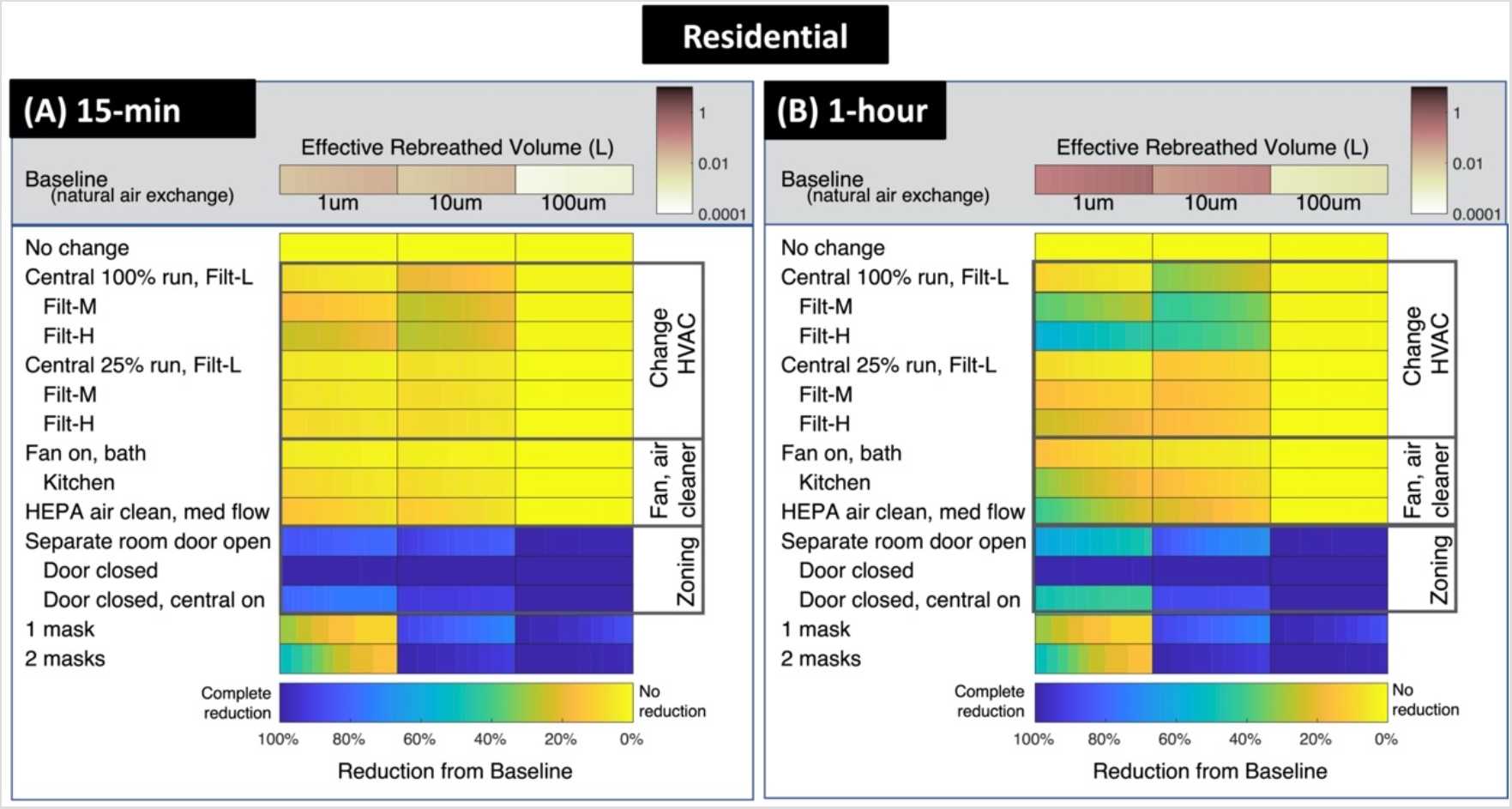
Baseline ERBV and mitigation cases for the residential case. 15-minute and 1-hour interactions are shown; 4-hour interactions are in the text, Fig. 3. In the 15-minute case, ERBV is low because accumulation has not yet occurred, and reductions are also low.

**Figure S.4.**
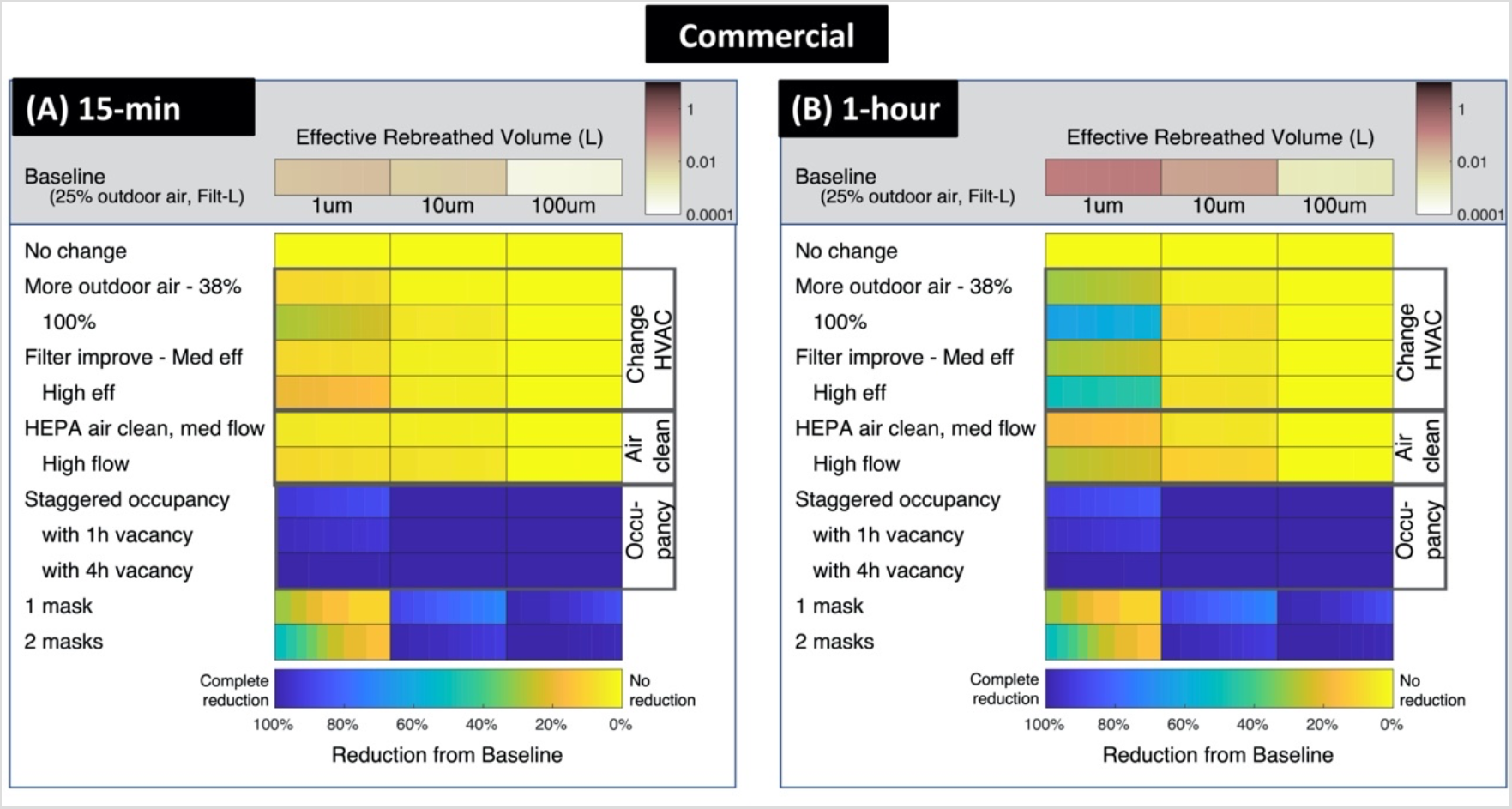
Baseline ERBV and mitigation cases for the commercial case. 15-minute and 1-hour interactions are shown; 4-hour interactions are in the text, Fig. 3.

**Figure S.5.**
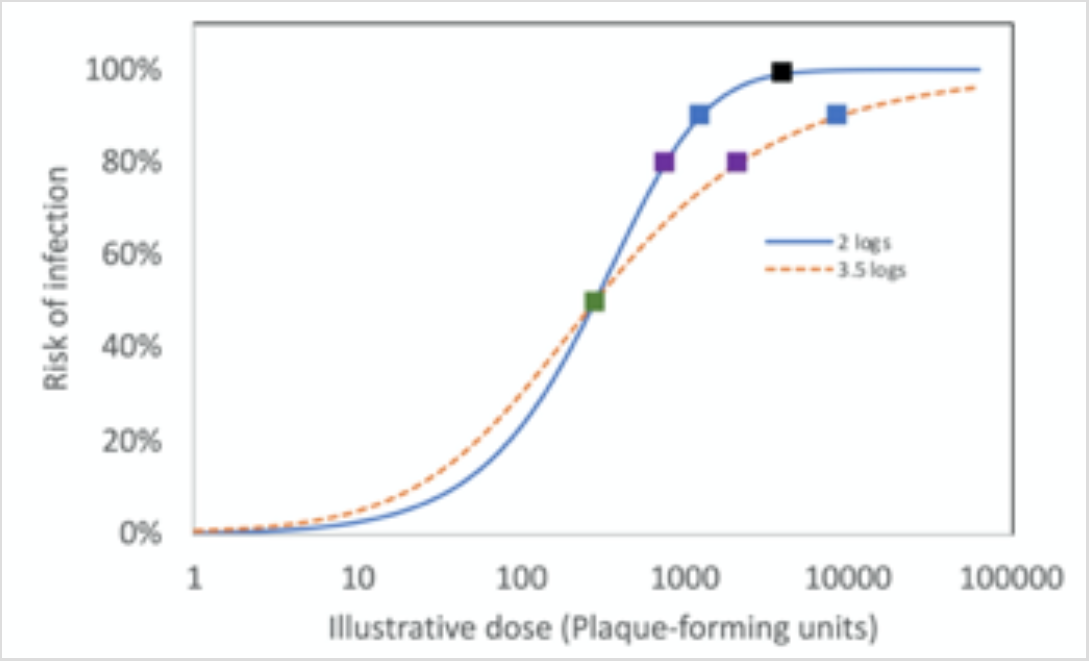
Illustrative dose-response curves with two widths. “Logs” indicates the base-10 logarithm of the ratio between the doses at 95% and 5% risk. Square markers indicate starting points for risk response to dose in Figure 4 (text). Dose-response curves are for illustration only and have not been determined for SARS-CoV-2.

**Figure S.6.**
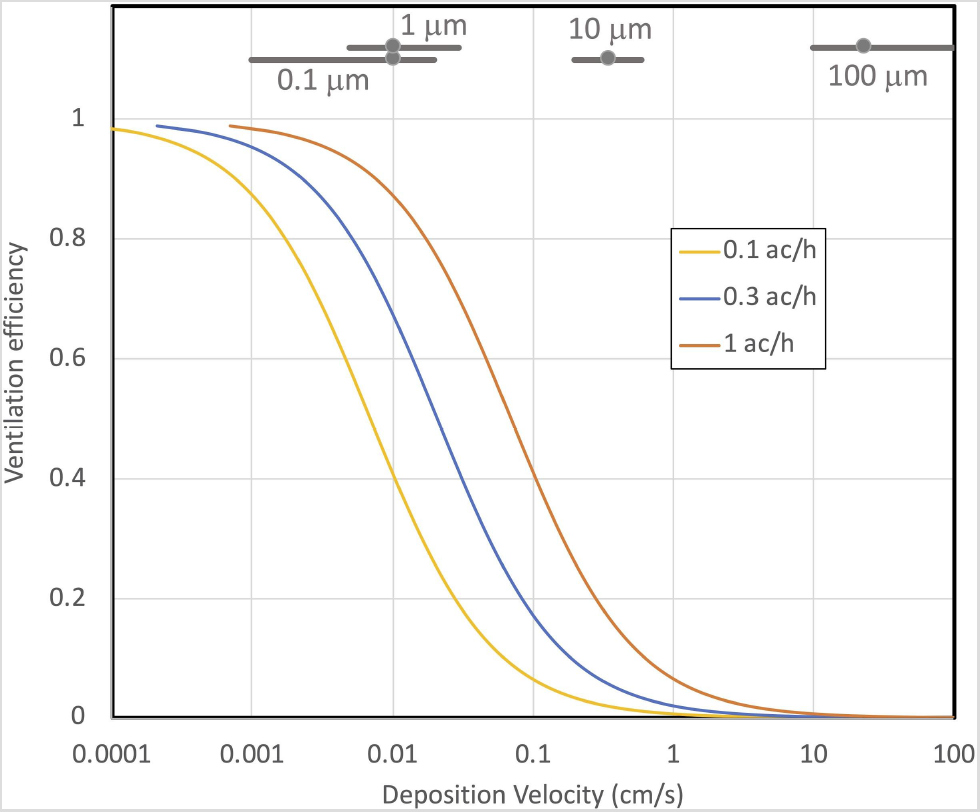
Ventilation efficiency for a range of deposition velocities. Ventilation efficiency is defined in Equation S.19.

## Tables

**Table S.1.**
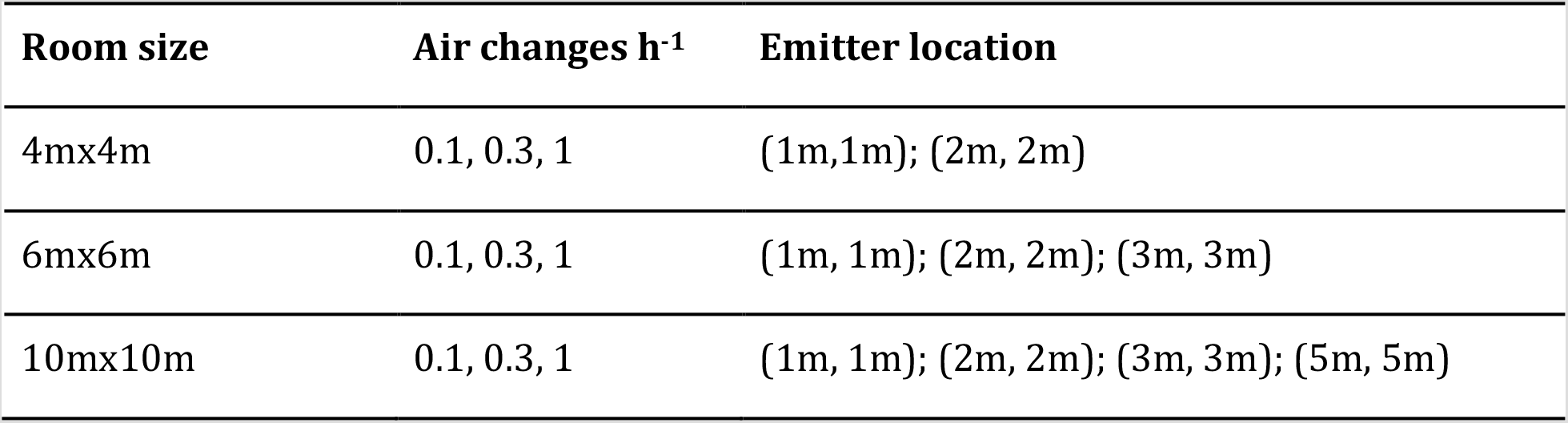
Inputs to indoor point-release simulations.

| Room size | Air changes h<sup>-1</sup> | Emitter location |
| --- | --- | --- |
| 4mx4m | 0.1, 0.3, 1 | (1m,1m); (2m, 2m) |
| 6mx6m | 0.1, 0.3, 1 | (1m, 1m); (2m, 2m); (3m, 3m) |
| 10mx10m | 0.1, 0.3, 1 | (1m, 1m); (2m, 2m); (3m, 3m); (5m, 5m) |

**Table S.2.** Deposition velocities

| Particle Diameter<br>(um) | Outdoor v<sub>d</sub><br>(m/s) | Indoor v<sub>d</sub> (m/s) |  | Indoor loss<br>rate<sup>a</sup> (h<sup>-1</sup>) |
| --- | --- | --- | --- | --- |
|  |  | Central value | Range |  |
| 0.1 | 3x10<sup>-7</sup> | 1x10<sup>-4</sup> | 0.1-2x10<sup>-4</sup> | 0.01-0.29 |
| 1 | 3x10<sup>-5</sup> | 1x10<sup>-4</sup> | 0.5-3x10<sup>-4</sup> | 0.072-0.44 |
| 10 | 3x10<sup>-3</sup> | 3.5x10<sup>-3</sup> | 2-6x10<sup>-3</sup> | 2.9-8.6 |
| 100 | 3x10<sup>-1</sup> | 0.23 | 0.1-1.0 | 140-1400 |

<sup>a</sup> For effective height of 2.5 m and deposition to upward-facing surfaces only.

**Table S.3.** Residential baseline values

| **Parameter** | **Value (range)** | **Justification** |
| --- | --- | --- |
| Floor area | 100-240 m<sup>2</sup> | Range of homes in USA |
| Ceiling Height | 2.5 m | Typical height |
| Air change rate | 0.1-1 ac h<sup>-1</sup> | Tight new homes to average older homes |

**Table S.4.** Residential mitigation cases. Discussion comments relate to 4-hour interactions [Figure 3A in text.]

| **Label** | **Detail** | **Support** | **Discussion** |
| --- | --- | --- | --- |
| Central air<br>100% run, Filt-L | 800-1600 cfm<br>[380 - 760 L s<sup>-1</sup>]<br>MERV 6 | a, b | Reduces ERBV<sub>10</sub> by about 50%; little effect on ERBV<sub>1</sub> because MERV6 filters are not rated to remove that size range |
| ", Filt-M | Same flow<br>MERV 11 | c | Reduces both ERBV<sub>1</sub> and ERBV<sub>10</sub> due to increase in filter quality. |
| ", Filt-H | Same flow<br>MERV 13 | d | Greater reduction in ERBV<sub>1</sub> than MERV11; however, MERV13 filters have a higher pressure drop, which is too great for some residential air handlers |
| Central air 25%<br>run, Filt-L | Same flow,<br>25% run time<br>MERV 6 | a, b, e | Compared with 100% run, less reduction because less air goes through the filter. 3-12% change in ERBV<sub>1</sub>; about 10-17% reduction in ERBV<sub>10</sub>. May correspond to normal operation during heating or cooling. |
| ", Filt-M | Same condition,<br>MERV 11 | c, e | Some effect on ERBV<sub>1</sub>. Compared with MERV 6, little change in ERBV<sub>10</sub>, as even the lower-rated filters remove this size range. |
| ", Filt-H | Same condition,<br>MERV 13 | d, e | About 30-50% reduction in ERBV<sub>1</sub>; similar ERBV<sub>10</sub> to Filt-M. |
| Bath fan | 80 cfm<br>[38 L s<sup>-1</sup>] | f | Lower end of mechanical ventilation. Reduces ERBV<sub>1</sub> by 13-40%. ERBV<sub>10</sub> is reduced by less than 10% because ventilation rate is lower than deposition rate. |
| Kitchen fan | 200 cfm<br>[94 L s<sup>-1</sup>] | g | Upper end of mechanical ventilation. Reduces ERBV<sub>1</sub> particles substantially (30-60%) and ERBV<sub>10</sub> somewhat (10-20%). |
| HEPA air clean,<br>med. flow | 200 cfm<br>[95 L s<sup>-1</sup>] | h | Portable unit that filters indoor air without changing flow into or out of the home. Requires HEPA rated filter. Substantial reduction in ERBV<sub>1</sub> and some reduction in ERBV<sub>10</sub>. Lower flow or filter of lower quality would not produce these reductions. |

Table S.4 (continued)
| Label | Detail | Support | Discussion |
| --- | --- | --- | --- |
| ", door closed | 0.6 cfm<br>[0.3 L s<sup>-1</sup>]<br>between rooms | i, j | Separating zones provides the greatest decrease in ERBV<sub>1</sub>, ERBV<sub>10</sub>, and ERBV<sub>100</sub> of all measures. Preventing release into a mixed space is more effective than removal. |
| ", door closed, central on | Same flow<br>between rooms,<br>central air<br>handler 100%<br>run | i, a, b | Central air handler with standard filter installed largely overcomes effect of closing doors |

Support notes for Table S.4.

**Table S.5.** Commercial baseline values

| **Parameter** | **Value (range)** | **Justification** |
| --- | --- | --- |
| Floor area | 150-200 m<sup>2</sup> | Open office arrangement, similar to residential |
| Ceiling height | 3 m | Typical height |
| Flow through air handler | 1600 cfm<br>[760 L/s] | Cooling requirement for the space |
| Outdoor air supply | 200 cfm<br>[94 L/s] | Minimum per standard |
| Filtration | MERV 8 | Minimum per standard |

**Table S.6.**
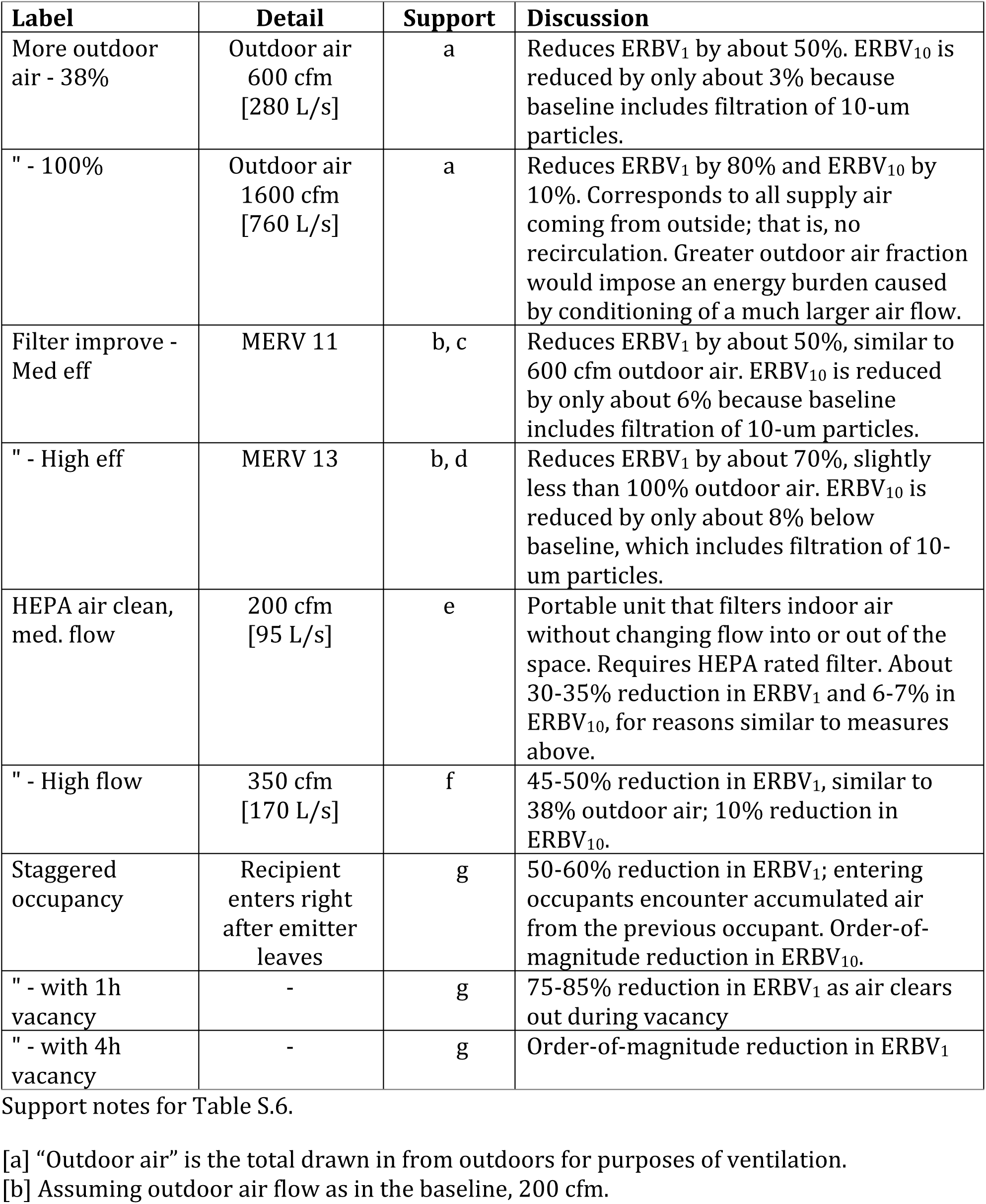
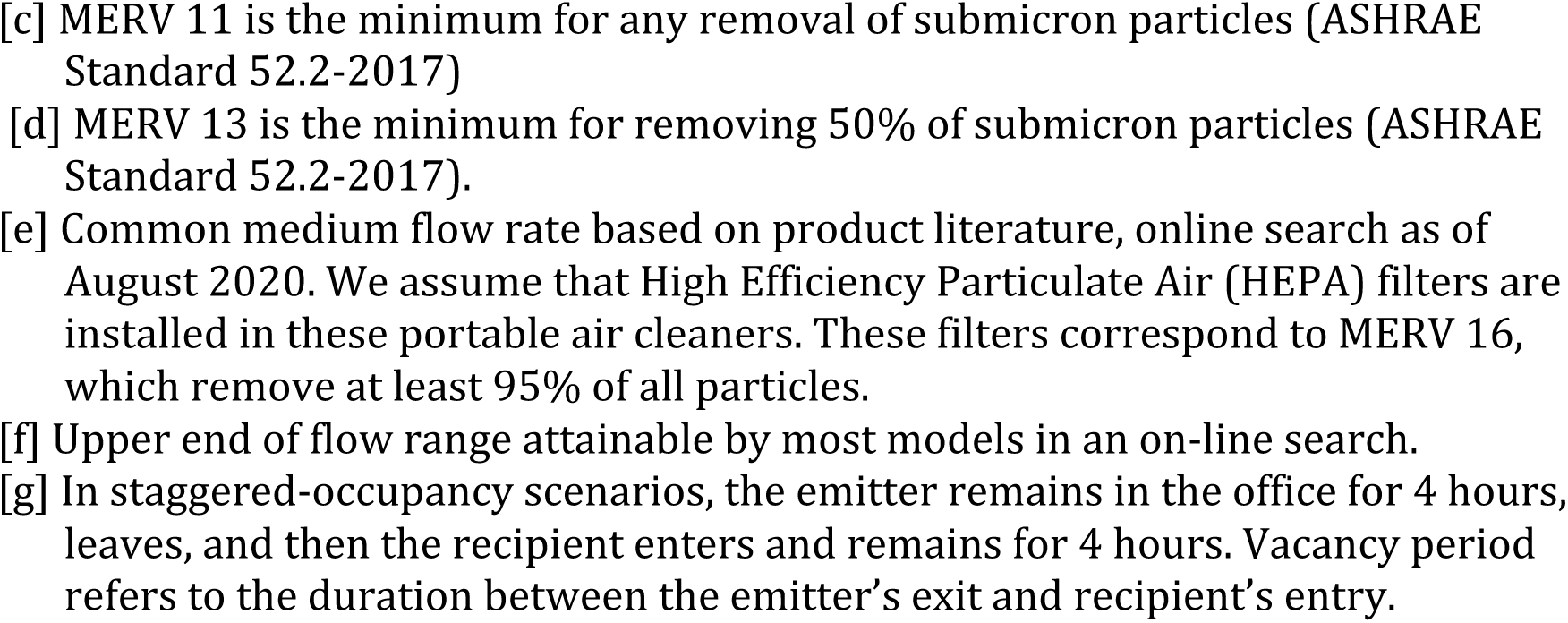
Commercial mitigation cases. Discussion comments relate to 4-hour interactions [Figure 3B in text.]

**Table S.7.**
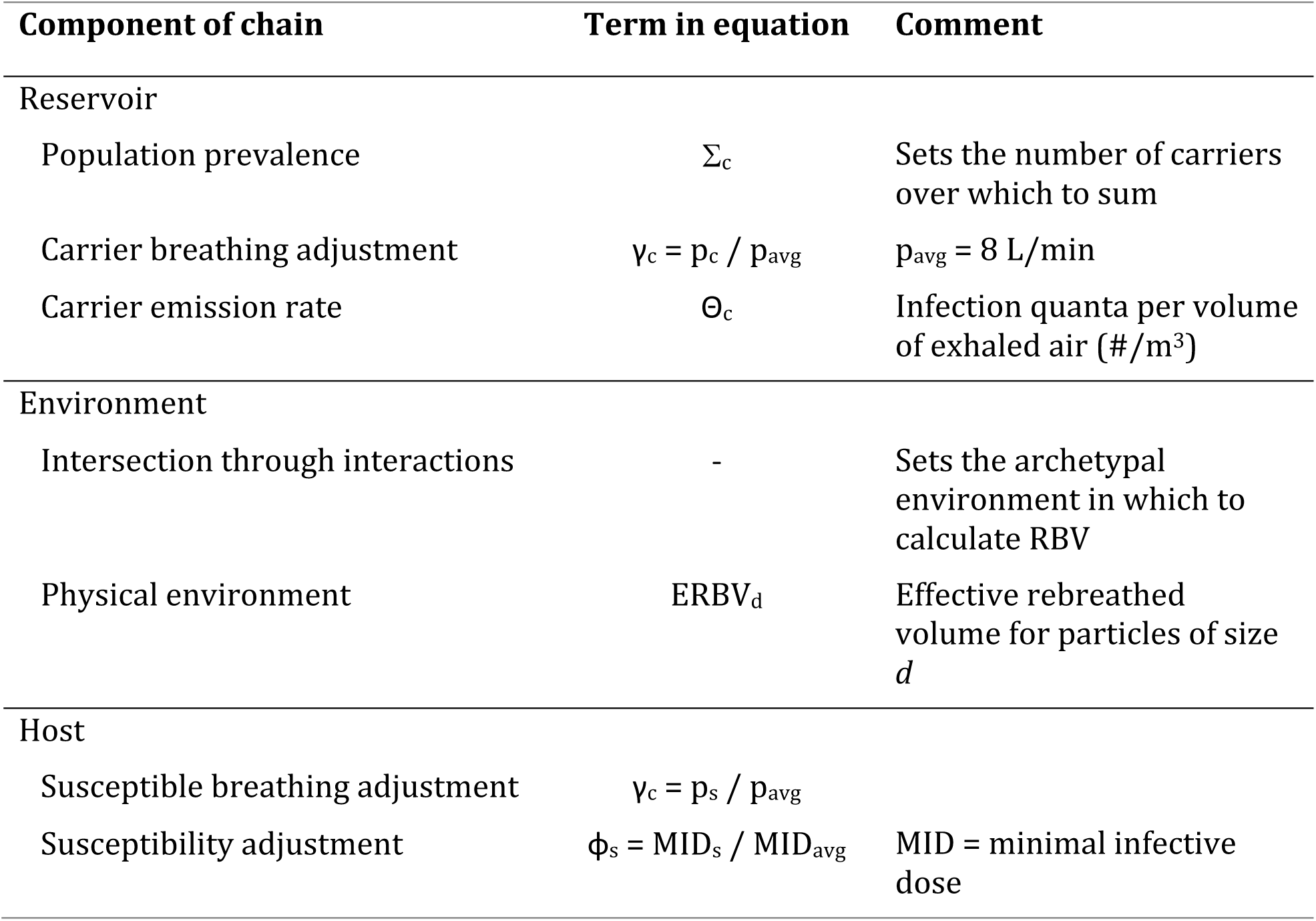
Terms in Wells-Riley equation for mean infection rate and their relationship to the chain of infection.

